# Modeling Monkeypox Epidemics: Thresholds, Temporal Dynamics, and Waning Immunity from Smallpox Vaccination”

**DOI:** 10.1101/2025.01.30.25321258

**Authors:** Samaila Jackson Yaga

## Abstract

This study investigates the dynamics of Monkeypox virus (MPXV) through a novel theoretical framework that extends classical epidemic threshold theory. The dual threshold theory is introduced, highlighting the interplay between the time-dependent basic reproduction number and the susceptible population density. Epidemic initiation is shown to occur when the time dependent reproductive number is greater than the threshold value of one and the susceptible population density at any time is greater than the critical threshold density of susceptibles. The model incorporates waning immunity from prior smallpox vaccination and immunity loss from previous MPXV infections, revealing complex epidemic behaviors such as oscillatory waves, prolonged outbreaks, and extended inter-epidemic periods under high transmission scenarios. Sensitivity analyses identify key drivers of epidemic initiation and progression, emphasizing the critical influence of waning immunity and zoonotic reservoirs. Public health implications underline the importance of targeted vaccination campaigns, rodent control, and continuous surveillance to reduce epidemic risks and prevent resurgence. This study provides actionable insights for managing MPXV outbreaks, while the dual threshold framework offers a robust theoretical foundation for understanding the dynamics of waning of vaccine cross immunity and zoonotic diseases.

## 1 Introduction

Monkeypox is a zoonotic disease caused by the monkeypox virus (MPXV), a member of the *Orthopoxvirus* genus in the *Poxviridae* family (WHO, 2024). It shares close biological and genetic similarities with *variola virus* (responsible for smallpox) (Cann et al., 2013; Rampogu et al., 2023).

Other notable members of this genus include cowpox virus and vaccinia virus. MPXV is classified as an emerging infectious disease (WHO, 2024; CDC, 2024a), due to its increasing prevalence and zoonotic potential (Islam et al., 2023).

MPXV is a double-stranded DNA virus with a brick-shaped morphology (Lu et al., 2023). The virus has two distinct clades: clade I (Congo Basin clade) and clade II (West African clade). Clade I is highly virulent, with a case-fatality ratio exceeding 10%, while clade II is less virulent, with a case-fatality ratio of less than 1% (Nakazawa et al., 2015; Wang et al., 2023). The first human case of monkeypox was reported in 1970, with subsequent outbreaks predominantly occurring in African countries such as the Democratic Republic of Congo, Nigeria, Cameroon, Gabon, and Liberia (Ghosh et al., 2023; Rampogu et al., 2023). The WHO declared monkeypox a global health emergency in 2023, highlighting its epidemic potential (Rampogu et al., 2023).

The transmission of MPXV can occur from animals to humans or between humans. Known reservoirs include rodents (e.g. squirrels, Gambian pouched rats, and dormice) and nonhuman primates (Wang et al., 2023). Animal-to-human transmission typically occurs through direct contact with infected animals’ excretions or secretions. Human-to-human transmission occurs through respiratory droplets, direct contact with contaminated materials (fomites), vertical transmission (CDC, 2024b), and sexual contact (Islam et al., 2023) Vulnerable populations, such as pregnant women, newborns, children, and immunocompromised individuals, are at greater risk of severe illness or death from MPXV infection (Wang et al., 2023).

Smallpox vaccination, widely administered during global eradication campaigns, has been shown to provide cross-protection against MPXV (Bunge et al., 2022). The vaccine’s efficacy in preventing monkeypox is approximately 85% (Lum et al., 2022). This cross-immunity arises due to genetic and antigenic similarities between *variola virus* and MPXV (Cann et al., 2013; Rampogu et al., 2023). However, following the eradication of smallpox, routine smallpox vaccination was discontinued worldwide in the late 20th century, leading to a gradual decline in immunity across populations. The protection conferred by smallpox vaccination is not lifelong, with immunity waning after approximately 3-5 years and continuing to decline over subsequent decades (CDC, 2024a). This waning immunity has been hypothesized as a significant driver of the resurgence of monkeypox outbreaks, particularly in regions with limited vaccination coverage (Banuet-Martinez et al., 2023).

In addition to waning immunity, factors such as increasing human-animal contact, deforestation, and population growth have amplified the risk of zoonotic spillover events. As immunity from historical smallpox vaccination campaigns continues to diminish, the population’s susceptibility to MPXV is expected to rise. This study aims to explore the impact of immunity decay on MPXV transmission dynamics, emphasizing the role of cross-protection from smallpox vaccination in mitigating the spread of the virus. Mathematical modeling provides a valuable framework for quantifying these dynamics and informing public health interventions.

## 2 Transmission Dynamics and Clinical Progression of MPXV

The transmission cycle illustrated in Figure 1 outlines the process of monkeypox virus (MPXV) transmission within human populations. Rodents are the primary reservoirs and known carriers of MPXV. Following exposure to the virus, an infected rodent undergoes an incubation period, progresses to an infectious state, and eventually recovers from the disease. In humans, after exposure to MPXV, the incubation period typically ranges between 2 and 21 days (WHO, 2024). The clinical progression of monkeypox disease is divided into three distinct phases, as shown in Figure 2:

**Fig. 1:**
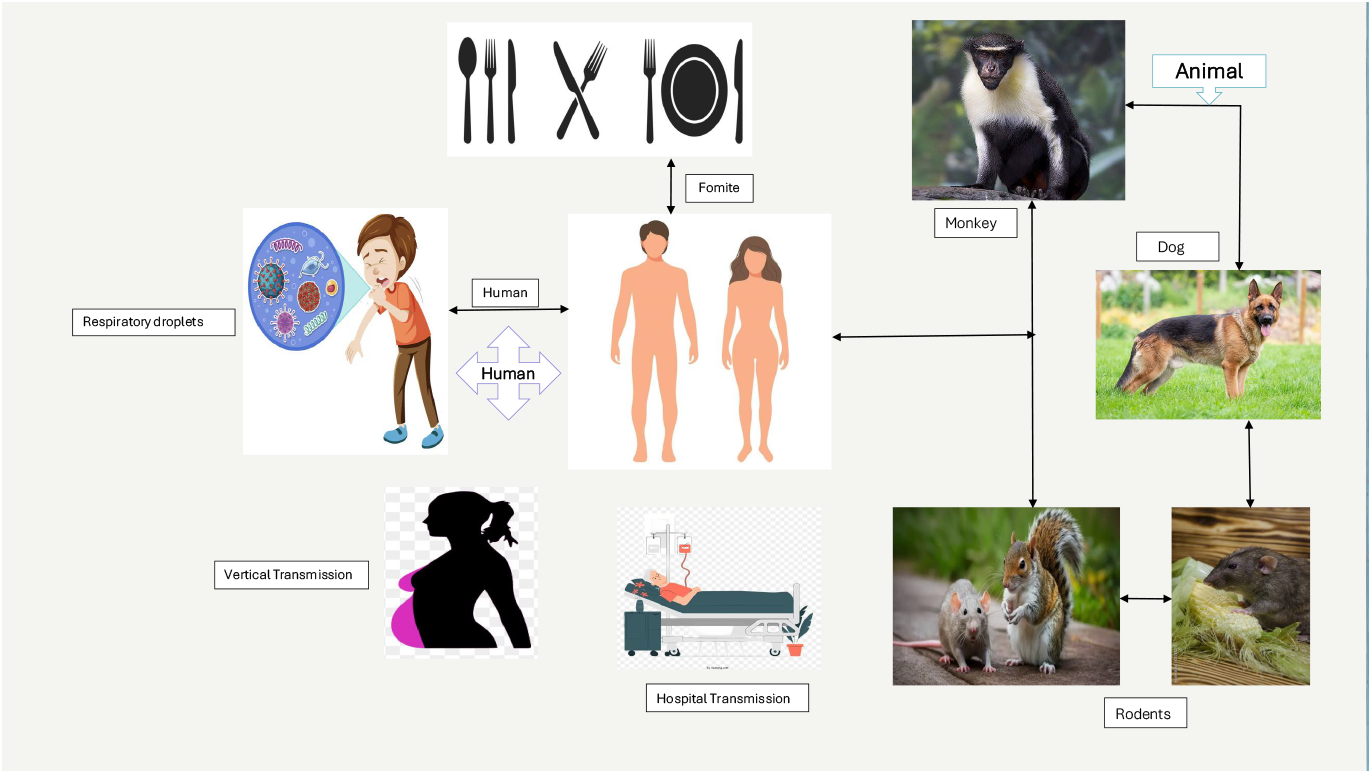
Showing the various mode of transmission of Monkeypox virus in both human and animal population

**Fig. 2:**
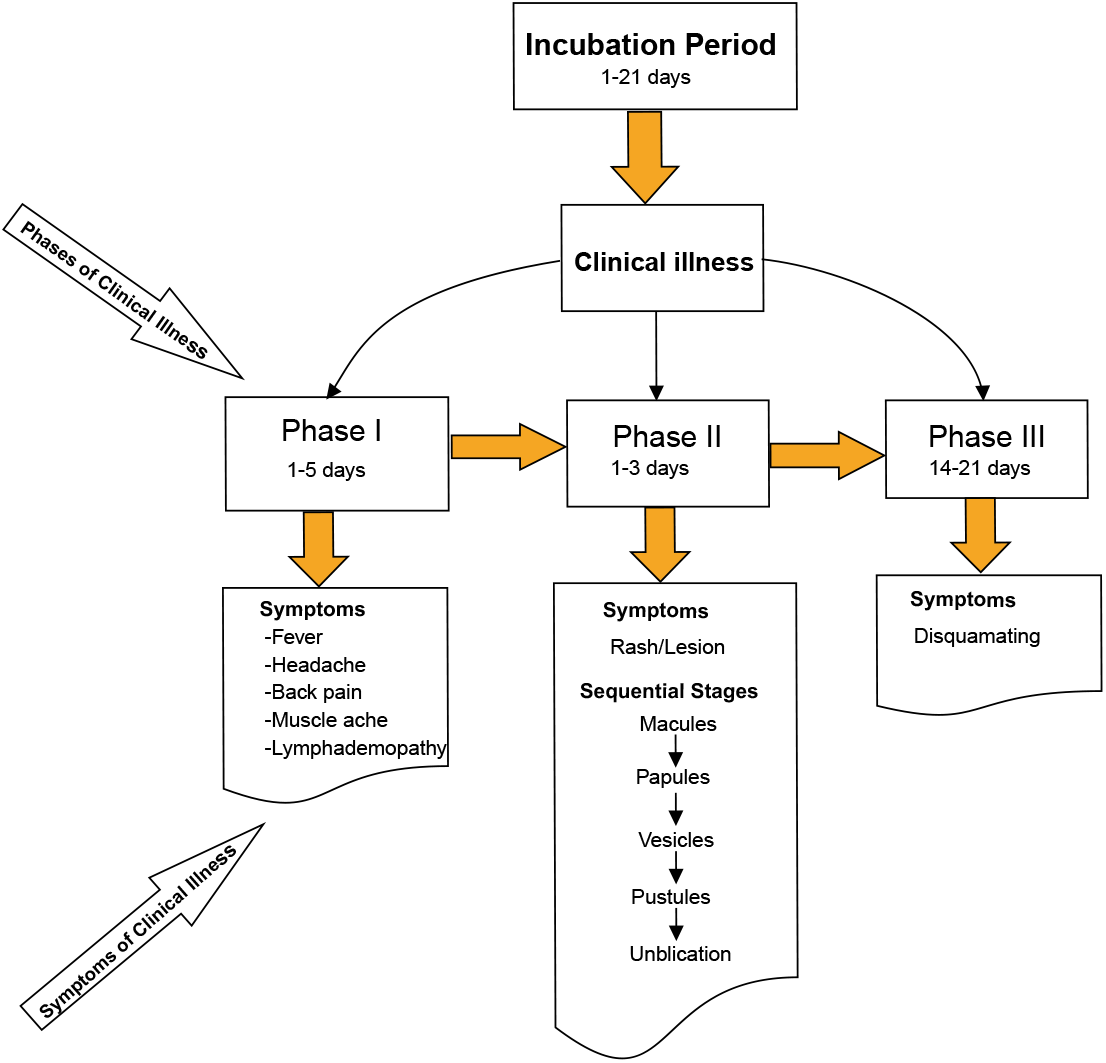
This is a wide figure. This is an example of a long caption that will wrap around. The caption is long and should be appropriately formatted. You can provide more details here as needed.

i. **Phase I**: This is the initial clinical stage, lasting 1 to 5 days. Symptoms during this phase include fever, headache, back pain, muscle ache, fatigue, and lymphadenopathy (swollen lymph nodes), which is a hallmark feature distinguishing MPXV infection from other poxvirus diseases.
ii. **Phase II**: Lasting 1 to 3 days, this phase is characterized by the appearance of a rash that progresses through sequential stages. The stages begin with macules, followed by papules, vesicles, pustules, and eventually umbilication.
iii. **Phase III**: This phase lasts 14 to 21 days and involves crusting of lesions, leading to desquamation (shedding of scabs). During this period, the individual remains infectious from the onset of symptoms until all scabs have fallen off.

Some cases of monkeypox can progress to severe and life-threatening conditions, necessitating hospitalization. Severe outcomes are predominantly associated with clade I MPXV, which is more virulent. In contrast, clade IIb is generally self-limiting, and most individuals recover within a few weeks (WHO, 2024; Wang et al., 2023; Rampogu et al., 2023) Un vaccinated individuals are at a significantly higher risk of severe complications, with sequelae observed in approximately 74% of unvaccinated cases compared to vaccinated populations (McCollum and Damon, 2014). Furthermore, individuals who recover from MPXV infection can still be reinfected (Li et al., 2024)

The eradication of smallpox, achieved through widespread vaccination campaigns, indirectly provided cross-protection against monkeypox due to the immunological similarities between the two viruses (Grant et al., 2020; WHO, 2024). However investigation into the role played by the cessation of routine smallpox vaccination that led to a decline in population immunity, leaving many individuals susceptible to MPXV is very important (Al-Tammemi et al., 2022).

In addition to human-to-human transmission via respiratory droplets, direct contact, or contaminated materials (fomites), vertical transmission (from mother to child) has also been documented (Wang et al., 2023; Dashraath et al., 2022). Individuals vaccinated against smallpox enter an immune compartment, which contributes to temporary protection against MPXV. Figure 2 illustrates the clinical state of the disease progression.

A framework for modeling the epidemiology of MPXV has been developed, primarily focusing on human-to-human and animal-to-human transmission pathways. The earliest model, introduced by Jezek et al. (1987), employed a stochastic approach to study MPXV transmission dynamics. This foundational work was further developed by Bhunu and Mushayabasa (2011), Betti et al. (2023), Peter et al. (2022), and Xiridou et al. (2024), with most models adopting compartmental approaches to explore the transmission mechanisms.

Several models have incorporated control strategies, aiming to evaluate interventions for mitigating MPXV spread. These include the works of Gani and Leach (2001), Usman et al. (2017), Emeka et al. (2018), Bankuru et al. (2020), Grant et al. (2020), and Yuan et al. (2022, 2023). More recent efforts, such as those by Clay et al. (2023), Awoke et al. (2024), and Rychtář et al. (2025), continue to refine these models to account for evolving epidemiological patterns.

Innovative approaches include the stochastic model developed by Khan et al. (2022), which incorporates environmental factors into transmission dynamics. Additionally, studies investigating coinfection with HIV, such as those by Bhunu et al. (2012) and Omame et al. (2024), highlight the interplay between MPXV and other infectious diseases.

Most of these models adopt the SEIR framework, with compartments representing Susceptible (S), Exposed (E), Infectious (I), and Recovered (R) individuals. Some models extend this framework by including additional compartments for vaccinated individuals and animal populations, such as rodents and non-human primates, to capture the zoonotic nature of MPXV transmission.

In this study, particular attention is given to population immunity stemming from prior smallpox vaccination. This is achieved by incorporating a compartment representing vaccinated and immune individuals. Additionally, the model accounts for the relative infectiousness of rodents in transmitting MPXV to humans compared to human-to-human transmission and incorporates reduced infectivity rates for hospitalized individuals relative to non-hospitalized infectious cases. These extensions represent novel contributions to the field.

### 2.1 Theoretical Model

The transmission dynamics of the monkeypox virus (MPXV) within a human population are modeled by incorporating the mechanisms of exposure via both rodent and human contact, as well as the influence of immunity resulting from prior smallpox vaccination. This theoretical framework focuses on the interactions between these compartments, with an emphasis on how immunity modulates the transmission process.

Let *S*_*m*_ denote the susceptible humans, who acquire MPXV through direct contact with infectious rodents, *I*_*r*_, at a rate *β*_*r*_, or through interaction with infectious humans, *I*_*m*_, at a rate *β*_*m*_, and with hospitalized individuals, *H*_*m*_, at a rate *αβ*. The parameter *α* accounts for the reduced infectivity of hospitalized individuals compared to those not hospitalized. Upon exposure to the virus, susceptible individuals transition to the exposed compartment, *E*_*m*_, where they undergo an incubation period and eventually develop symptoms. These exposed individuals become infectious at a rate *θ*_*m*_. A fraction *p* of infectious individuals are hospitalized, reflecting the severity of the disease, at the rate *ϕ*_*m*_. The remaining proportion, (1 − *p*), recover and acquire temporary immunity to MPXV at a rate *γ*_*m*_.Infectious individuals and hospitalized may die due to the severity of the disease at rates *δ*_1_ and *δ*_2_, respectively. Additionally, vertical transmission of MPXV is incorporated into the model, where a proportion *τ* of newborns transition to the susceptible compartment at a rate *b*_*h*_, while the remaining fraction, (1 − *τ*), are exposed to infectious, also at a rate *b*_*h*_. All compartments are subject to death rates, *μ*, reflecting the turnover of the population.

This setup allows us to focus strictly on the effects of waning immunity from smallpox vaccination on the dynamics of monkeypox transmission. By incorporating the dynamics of *R*_*s*_ and *S*_*m*_. The model captures the gradual reduction in population-level immunity and the corresponding increase in susceptibility to monkeypox. This approach highlights the long-term epidemiological implications of historical vaccination campaigns and their cessation after smallpox eradication.

This model explicitly examines the impact of smallpox vaccination-induced immunity on the transmission dynamics of monkeypox, emphasizing the role of immunity waning over time. The force of infection due to MPXV among humans, denoted by, *λ*_*h*_, determines the rate at which susceptible humans are infected when they make an effective contact with an infectious human. This is given by

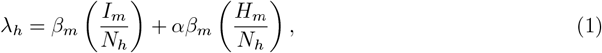

where *β*_*m*_ is the human-to-human transmission rate, and *α* accounts for reduced infectivity from hospitalized individuals (*H*_*m*_).

The force of infection from animal to human, denoted *λ*_*r*_, determines the rate at which susceptible humans are infected through contact with infectious animals:

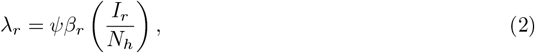

where *ψ* represents the relative infectivity of infectious rodents (*I*_*r*_) to humans compared to human-to-human transmission.

The total force of infection against humans due to MPXV is the sum of human-to-human (equation (**??**) and rodent-to-human pathways (equation **??**):

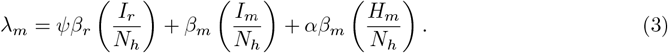

#### Assumptions and Model Setup for Smallpox

Since smallpox has been eradicated globally, routine smallpox vaccination programs have ceased. Therefore, in this model, we set the smallpox vaccination rate *v* = 0. Furthermore, we assume that there are no actively susceptible (*S*_*s*_ = 0), exposed (*E*_*s*_ = 0), or infectious (*I*_*s*_ = 0) individuals for smallpox. However, we define (*R*_*s*_ = *n*), where *n* represents the number of individuals who were vaccinated against smallpox and are initially fully immune. Although smallpox has been eradicated, we assume that children born into the population who are susceptible to monkeypox would also have been susceptible to smallpox in the absence of vaccination. Consequently, all newborn susceptibles move directly into the monkeypox-susceptible compartment (*S*_*m*_) at a rate of *τb*_*h*_, where *τ* is the proportion of newborns that are susceptible, and *b*_*h*_ is the human birth rate.

While smallpox itself is not part of the model’s active transmission dynamics, we are interested in the population that initially received the smallpox vaccine (*R*_*s*_) and subsequently lost immunity over time. The immunity in this vaccinated population wanes at a rate *σ*, transitioning individuals from the *R*_*s*_ compartment to the monkeypox-susceptible compartment, *S*_*m*_. It is important to note that the only smallpox-related compartment that changes dynamically over time is the vaccinated immune compartment (*R*_*s*_). This change occurs exclusively due to the waning immunity effect, modeled by the term, which reduces while proportionally increasing .

A key feature of this model is the incorporation of two distinct sources of immunity

#### Immunity Due to Smallpox Vaccination

Individuals who were previously vaccinated against smallpox are assumed to transition to the immune compartment at a given rate . Individuals in the immune/Vaccinated compartment lose immunity back to susceptible class is satisfied by the differential equation

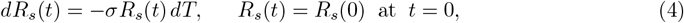

where *σ* represents the waning rate of smallpox vaccine-derived immunity. equation (4) can be integrated to yield:

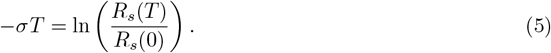

Here, *R*_*s*_(*T*) denotes the surviving immune at time *T*, while *σ* captures the rate at which immunity decreases in the vaccinated/immune population over time.

#### Immunity Due to Monkeypox Virus Infection

The second source of immunity arises from direct infection with the Monkeypox virus (MPXV). Recovered individuals (*R*_*m*_) who were previously infected with MPXV acquire temporary immunity. Over time, however, these individuals may lose their immunity at a rate, leading to a potential return to the susceptible compartment. This dynamic highlights the transient nature of immunity following natural infection.

The inclusion of these two immunity sources introduces a dual-layered perspective on population susceptibility. The waning immunity from both smallpox vaccination and monkeypox recovery contributes to an evolving susceptible population, which is crucial for understanding long-term transmission dynamics. The parameters *σ* and *σ*_*m*_ serve as key determinants of how quickly immunity is lost in their respective compartments, directly influencing the potential for future outbreaks.The transition diagram (Figure 3) and the system of ordinary differential equations that describes the transmission dynamics of the monkeypox virus (MPXV) within the human population are given in

**Fig. 3:**
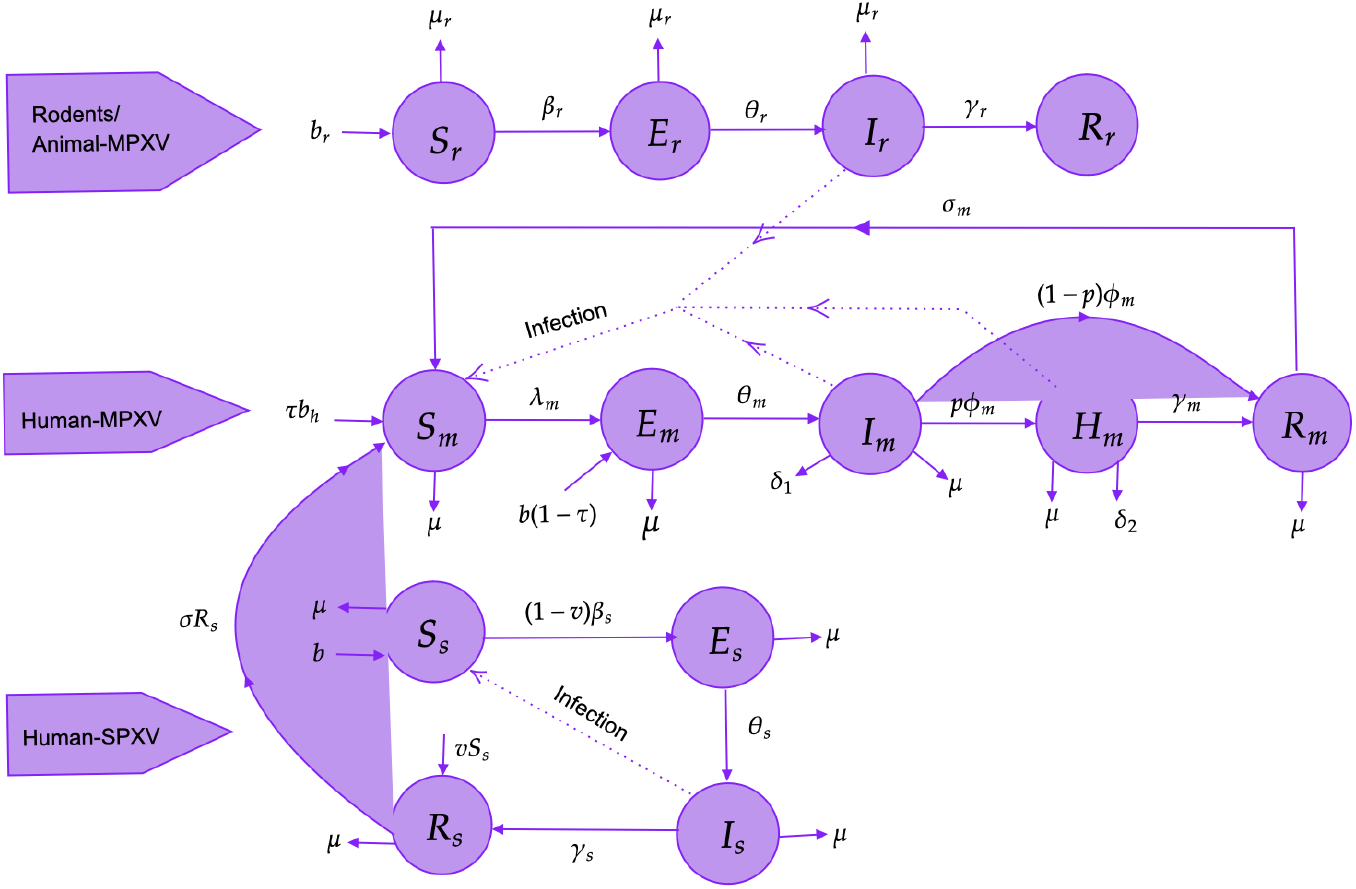
Transition diagram for Monkeypox virus transmission process

The system of differential equations governing MPXV dynamics in humans is given by:

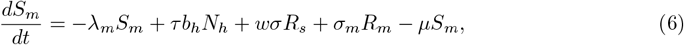

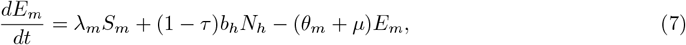

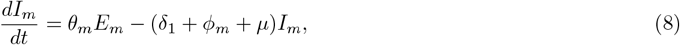

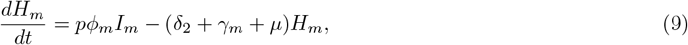

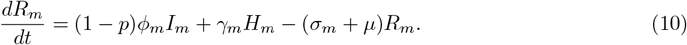

The dynamics for Smallpox are described as:

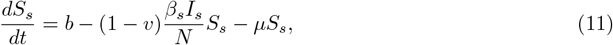

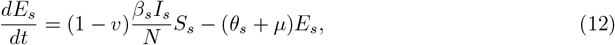

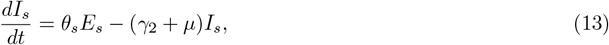

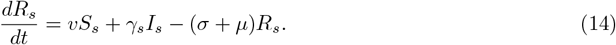

The total human population is conserved:

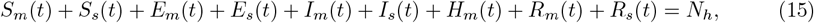

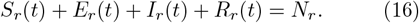

The dynamics for rodents are described as:

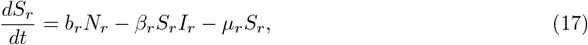

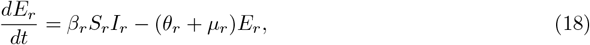

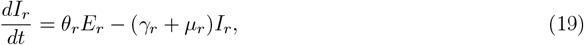

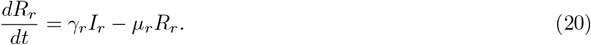

## 3 Impact of Immunity Decay on Susceptible Dynamics

The waning of immunity indirectly influences the dynamics of the population susceptible to the Monkeypox virus, denoted as *S*_*m*_(*t*). This influence is inherently temporal and manifests through the replenishment of the susceptible class. Specifically, individuals from the vaccinated class, represented by *R*_*s*_(*t*), transition back into the susceptible population due to the decay of immunity over time. This decay is attributed to the waning efficacy of the smallpox vaccine, which diminishes the longterm protection initially conferred by vaccination. To establish this theory mathematically into this phenomenon, The solution *S*_*m*_(*t*), from the system of equation in (6)-(10) is:

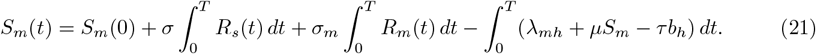

Here, *S*_*m*_(*t*) is replenished by three components, one from the population previously vaccinated with smallpox vaccine, that wanes at a rate *σ*, the second one is the rate of loss of temporary immunity *σ*_*m*_ for individuals who recovered from monkeypox disease. The third one is the birth of new susceptible *τb*_*h*_.

The waning rate *σ* increases or replenishes *S*_*m*_(*t*) leading to a large pool of susceptible. This in turn affect the dynamics of the susceptible (*S*_*m*_) through the component:

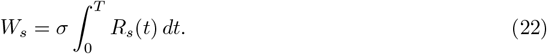

The immunity decay from loss of temporary immunity for individuals infected with monkeypox affect the size of *S*_*m*_(*t*) through

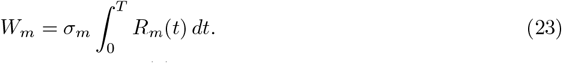

The solution to equation (14) is obtained in equation (5) as

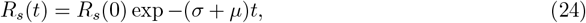

and the solution for (10) is obtained by integration to get

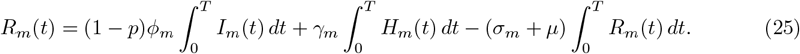

### 3.1 Basic Reproductive number(*R*_0_)

The basic reproduction number, *R*_0_, is a fundamental concept in epidemiology that quantifies the potential for a pathogen to spread within a population. It represents the average number of secondary infections produced by a single infected individual in a completely susceptible population. For the monkeypox virus (MPXV), *R*_0_ helps to assess transmission dynamics, considering human-to-human and rodent-to-human transmission. The Next Generation Matrix is given by 𝒢= 𝒯𝒱^−1^, and the spectral radius (largest eigenvalue) of 𝒢 represents the basic reproduction number. The Next Generation Matrix (NGM) is derived from the system of differential equation, focusing on the infectious compartments:

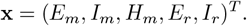

The transmission matrix is derived from the force of infection. This matrix is obtained from infected sub-system of (6)-(10) using the large domain(Diekmann et al., 2010; Van den Driessche, 2017; Yaga and Saporu, 2024b), hence the transmission matrix is given as:

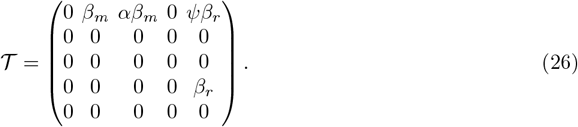

and the transition matrix of the infected sub-system 𝒱 is:

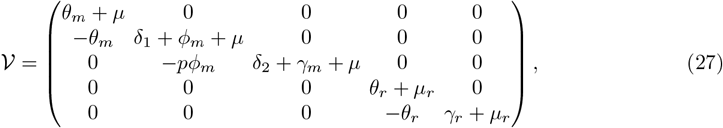

and its inverse is given as

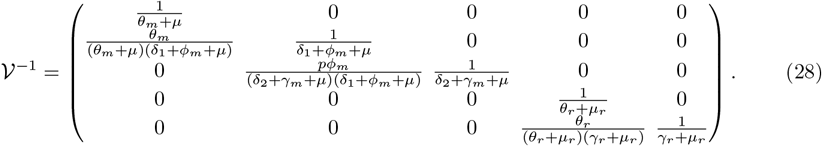

The basic reproduction number for the MPXV transmission model is expressed as:

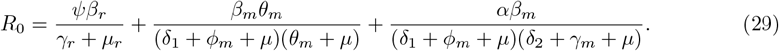

The total reproduction number *R*_0_ can be decomposed into two primary components representing different transmission pathways:

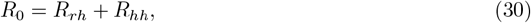

where:

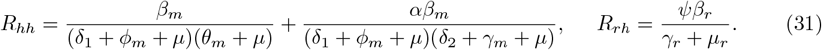

Here, *R*_*hh*_ represents the reproduction number due to human-to-human transmission, while *R*_*rh*_ represents the reproduction number attributed to the transmission of rodent-to-human spillover. These components are discussed in detail below.

### Human-to-Human Transmission (*R*_*hh*_)

The human-to-human transmission component is given by:

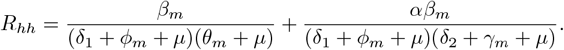

This expression shows that *R*_*hh*_ is directly proportional to the human-to-human transmission rate *β*_*m*_ and inversely proportional to the rates of disease progression (*ϕ*_*m*_, *θ*_*m*_), recovery (*γ*_*m*_) disease induce death (*δ*_1_, *δ*_1_) and natural death (*μ*). A slower progression or recovery process results in a higher *R*_*hh*_, indicating that infection persists longer within the human population, increasing the potential for sustained transmission.

### Rodent-to-Human Transmission (*R*_*rh*_)

The component of the transmission from rodent to human is given by:

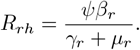

Here, *R*_*rh*_ depends on the transmission rate from rodent to human *β*_*r*_, the recovery rate of infected rodents (*γ*_*r*_) and the rate of death of rodents (*μ*_*r*_). The parameter *ψ* acts as a scaling factor, capturing the efficiency of rodent-to-human transmission. A high rodent population and increased contact between rodents and humans can lead to substantial *R*_*rh*_, emphasizing the role of zoonotic transmission in maintaining MPXV outbreaks.

Understanding the relative contributions of *R*_*hh*_ and *R*_*rh*_ provides valuable insights into transmission dynamics and prioritization of control measures. The basic reproduction number *R*_0_ serves as a key metric to assess the epidemic potential of MPXV:

i. *R*_0_ > 1: Each infected individual generates more than one secondary case, indicating the potential for sustained transmission and epidemic spread.
ii. *R*_0_ < 1: The infection cannot sustain itself in the population and will eventually die out.

Any control strategy that aims at reducing the overall potential of MPXV epidemic should lower that basic reproductive number below one (Max(*R*_*hh*_, *R*_*rh*_) < 1).

### 3.2 Temporal Evolution of *R*_0_

The time-dependent basic reproduction number, denoted as *R*_0_(*t*), represents the average number of secondary infections caused by an infectious individual at time *t* within a population. It incorporates temporal variations in population immunity, control measures, and epidemic risk, primarily through the indirect effects mediated by the susceptible population. For the model presented, the baseline basic reproduction number is expressed as:

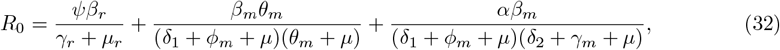

where the terms reflect contributions from different transmission pathways and demographic or epidemiological processes.The relationship between *R*_0_ and the susceptible population *S*_*m*_(*t*) facilitates the exploration of the impact of waning immunity on the susceptible pool through the temporal evolution of *R*_0_. The time-dependent reproduction number *R*_0_(*t*) is formulated as:

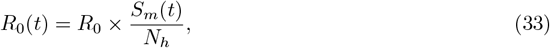

where *N*_*h*_ denotes the total human population. Substituting for *R*_0*m*_, the temporal evolution function becomes:

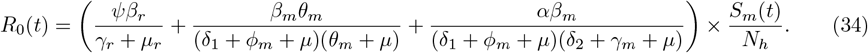

This formulation links the time evolution of *R*_0_ directly to changes in the susceptible population over time, allowing an analysis of how waning immunity influences transmission dynamics.

## 4 Sensitivity Analysis

Sensitivity analysis quantifies the impact of parameters on a response function (metric), such as the basic reproduction number (*R*_0_) (Yaga and Saporu, 2024b; Diekmann et al., 2013) or the probability of disease extinction (Yaga and Saporu, 2024a). Several methods exist for performing sensitivity analysis, including local sensitivity analysis (e.g., forward sensitivity index) and global sensitivity analysis (e.g., Latin Hypercube Sampling, LHS) (Blower and Dowlatabadi, 1994; Marino et al., 2008)

### 4.1 Local Sensitivity: Forward Sensitivity index

The forward sensitivity index of *R*_0_ with respect to for example, *ψ*, is define as:

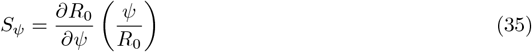

where *S*_*ψ*_ is dimensionless sensitivity index. When *S*_*ψ*_ > 0, it implies that *R*_0_ is increased when *ψ* is increased and when *S*_*ψ*_ < 0, *R*_0_ decreases when *ψ* is increased. Generally, |*S*_*ψ*_| is the magnitude of *S*_*ψ*_, that shows the strength of sensitivity of *R*_0_ on *S*_*ψ*_.This enable the identification of critical parameters, assess their impact, and refine interventions to better control disease transmission.

### 4.2 Global Sensitivity: Latin Hypercube Sampling (LHS)

Latin Hypercube Sampling (LHS) is a statistical technique designed to generate plausible samples from a multi-dimensional parameter space (Blower and Dowlatabadi, 1994). This method is widely used in theoretical epidemiology for global sensitivity analysis due to its efficiency in exploring parameter space with fewer samples than simple random sampling. In the LHS method, the distribution of each parameter, denoted by *ψ*_*v*_ for *v* = 1, 2, 3, …, *k*, is divided into equal probability intervals *I*_*vj*_, where:

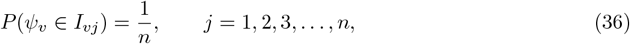

with *I*_*jv*_ representing the interval for parameter *ψ*_*j*_ within the range [*ψ*_*a*_, *ψ*_*b*_]. For the *v*^th^ sample value of *ψ*_*v*_, the expression is:

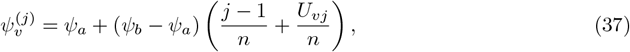

where *U*_*vj*_ ∼ *U* (0, 1) (uniformly distributed between 0 and 1), *j* = 1, 2, 3, …, *n*, and *n* denotes the number of samples. To complete the sampling, the intervals are permuted, and an LHS matrix *S*_*vj*_ is constructed, where each row corresponds to a sample and each column corresponds to a parameter. This matrix is subsequently used to run *n* simulations of the system of nonlinear differential equations, with parameters sampled according to *S*_*vj*_.

#### Discussion of Sensitivity Analysis

Table 3 present the sensitivity results for parameters of basic reproductive number (*R*_0_) and Figure 4 shows the trend and strength of influence exert by these parameters on *R*_0_. Here, a higher sensitivity index indicates a longer influence on the ability of disease spread in the population.

**Table 1:** Parameters for MPXV and SMPXV Transmission Model (Humans and Rodents)

| Param | Description | Value(s) | Reference |
| --- | --- | --- | --- |
| $\beta_m$ | Human-to-human transmission rate | 30 | Grant et al. (2020) |
| $\beta_r$ | Rodent transmission rate | 32.85 | Bankuru et al. (2020) |
| $\alpha$ | Reduced infectivity of hospitalized humans | 0.1 | Assumed |
| $\psi$ | Relative infectivity rate of rodent | 0 - 10 | Assumed |
| $\theta_m$ | Rate of exposed humans becoming infectious | 10-40 | Yuan et al. (2022) |
| $\delta_1$ | Rate at which infectious humans die due to MPXV | 28.8 | Assumed |
| $\delta_2$ | Rate at which hospitalized humans die due to MPXV | 28.8 | Assumed |
| $\phi_m$ | Disease progression rate for humans | 0.1 - 1 | Wang et al. (2023) |
| $\gamma_m$ | Recovery rate for hospitalized humans | 17.7 | Olivencia et al. (2024) |
| $\sigma_m$ | Immunity loss rate for recovered humans | 0-30 | Assumed |
| $\sigma$ | Immunity loss rate for recovered humans | 0 - 1 | Assumed |
| $w$ | Proportion of individuals in $R_s$ that lose immunity | 0 - 1 | Assumed |
| $p$ | Proportion of hospitalized infected humans | 0.1 - 1 | Assumed |
| $\mu$ | Natural mortality rate for humans | 0.01 - 0.05 | WHO (2024) |
| $b_h$ | Birth rate in human population | 0.01 - 0.05 | WHO (2024) |
| $\tau$ | The proportion of individuals born into $S_m$ | 0-1 | Assumed |
| $\beta_s$ | Vaccination transmission rate | 0.001 | Assumed |
| $\theta_s$ | Rate at which $E_s$ moves to $I_s$ for SPXV | 0.001 | Assumed |
| $\gamma_s$ | Rate of recovery from SPXV | 0.0001 | Assumed |
| $v$ | Proportion of population vaccinated | 0-1 | WHO (2024) |
| $\mu_r$ | Natural mortality rate for rodents | 0.1 - 0.5 | Assumed |
| $\gamma_r$ | Recovery rate for infected rodents | 17.7 | Assumed |
| $b_r$ | Birth rate of rodent | 0.2 | Bhunu and Garira (2009) |
| $\theta_r$ | Rate of exposed rodents becoming infectious | 13 | Assumed |

**Table 2:**
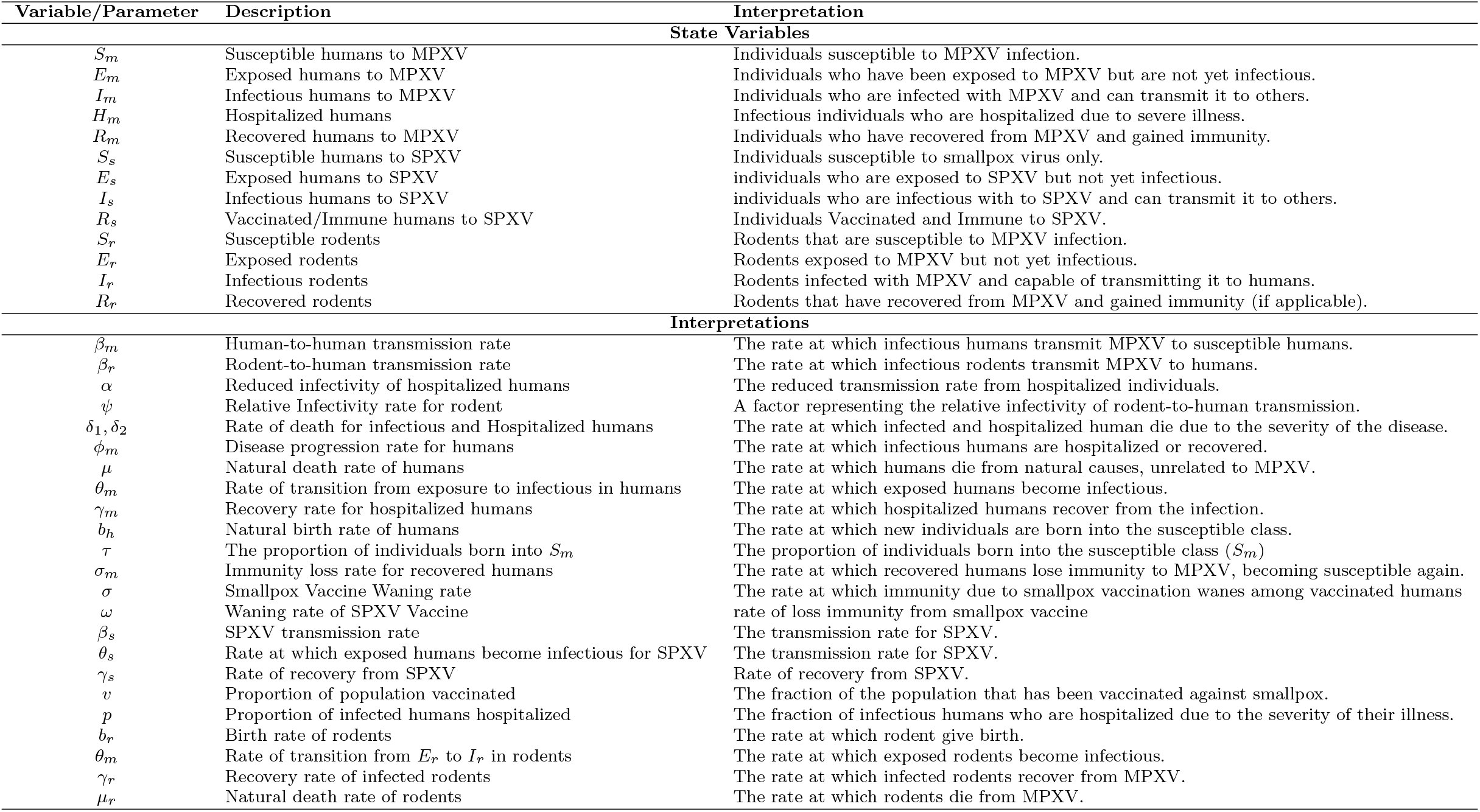
State Variables and Parameters of the MPXV Transmission Model.

| Variable/Parameter | Description | Interpretation |
| --- | --- | --- |
| **State Variables** |  |  |
| $S_m$ | Susceptible humans to MPXV | Individuals susceptible to MPXV infection. |
| $E_m$ | Exposed humans to MPXV | Individuals who have been exposed to MPXV but are not yet infectious. |
| $I_m$ | Infectious humans to MPXV | Individuals who are infected with MPXV and can transmit it to others. |
| $H_m$ | Hospitalized humans | Infectious individuals who are hospitalized due to severe illness. |
| $R_m$ | Recovered humans to MPXV | Individuals who have recovered from MPXV and gained immunity. |
| $S_s$ | Susceptible humans to SPXV | Individuals susceptible to smallpox virus only. |
| $E_s$ | Exposed humans to SPXV | Individuals who are exposed to SPXV but not yet infectious. |
| $I_s$ | Infectious humans to SPXV | Individuals who are infectious with to SPXV and can transmit it to others. |
| $R_s$ | Vaccinated/Immune humans to SPXV | Individuals Vaccinated and Immune to SPXV. |
| $S_r$ | Susceptible rodents | Rodents that are susceptible to MPXV infection. |
| $E_r$ | Exposed rodents | Rodents exposed to MPXV but not yet infectious. |
| $I_r$ | Infectious rodents | Rodents infected with MPXV and capable of transmitting it to humans. |
| $R_r$ | Recovered rodents | Rodents that have recovered from MPXV and gained immunity (if applicable). |
| **Interpretations** |  |  |
| $\beta_m$ | Human-to-human transmission rate | The rate at which infectious humans transmit MPXV to susceptible humans. |
| $\beta_r$ | Rodent-to-human transmission rate | The rate at which infectious rodents transmit MPXV to humans. |
| $\alpha$ | Reduced infectivity of hospitalized humans | The reduced transmission rate from hospitalized individuals. |
| $\psi$ | Relative Infectivity rate for rodent | A factor representing the relative infectivity of rodent-to-human transmission. |
| $\delta_1, \delta_2$ | Rate of death for infectious and Hospitalized humans | The rate at which infected and hospitalized human die due to the severity of the disease. |
| $\phi_m$ | Disease progression rate for humans | The rate at which infectious humans are hospitalized or recovered. |
| $\mu$ | Natural death rate of humans | The rate at which humans die from natural causes, unrelated to MPXV. |
| $\theta_m$ | Rate of transition from exposure to infectious in humans | The rate at which exposed humans become infectious. |
| $\gamma_m$ | Recovery rate for hospitalized humans | The rate at which hospitalized humans recover from the infection. |
| $b_h$ | Natural birth rate of humans | The rate at which new individuals are born into the susceptible class. |
| $\tau$ | The proportion of individuals born into $S_m$ | The proportion of individuals born into the susceptible class ($S_m$) |
| $\sigma_m$ | Immunity loss rate for recovered humans | The rate at which recovered humans lose immunity to MPXV, becoming susceptible again. |
| $\sigma$ | Smallpox Vaccine Waning rate | The rate at which immunity due to smallpox vaccination wanes among vaccinated humans |
| $\omega$ | Waning rate of SPXV Vaccine | rate of loss immunity from smallpox vaccine |
| $\beta_s$ | SPXV transmission rate | The transmission rate for SPXV. |
| $\theta_s$ | Rate at which exposed humans become infectious for SPXV | The transmission rate for SPXV. |
| $\gamma_s$ | Rate of recovery from SPXV | Rate of recovery from SPXV. |
| $v$ | Proportion of population vaccinated | The fraction of the population that has been vaccinated against smallpox. |
| $p$ | Proportion of infected humans hospitalized | The fraction of infectious humans who are hospitalized due to the severity of their illness. |
| $b_r$ | Birth rate of rodents | The rate at which rodent give birth. |
| $\theta_m$ | Rate of transition from $E_r$ to $I_r$ in rodents | The rate at which exposed rodents become infectious. |
| $\gamma_r$ | Recovery rate of infected rodents | The rate at which infected rodents recover from MPXV. |
| $\mu_r$ | Natural death rate of rodents | The rate at which rodents die from MPXV. |

**Table 3:**
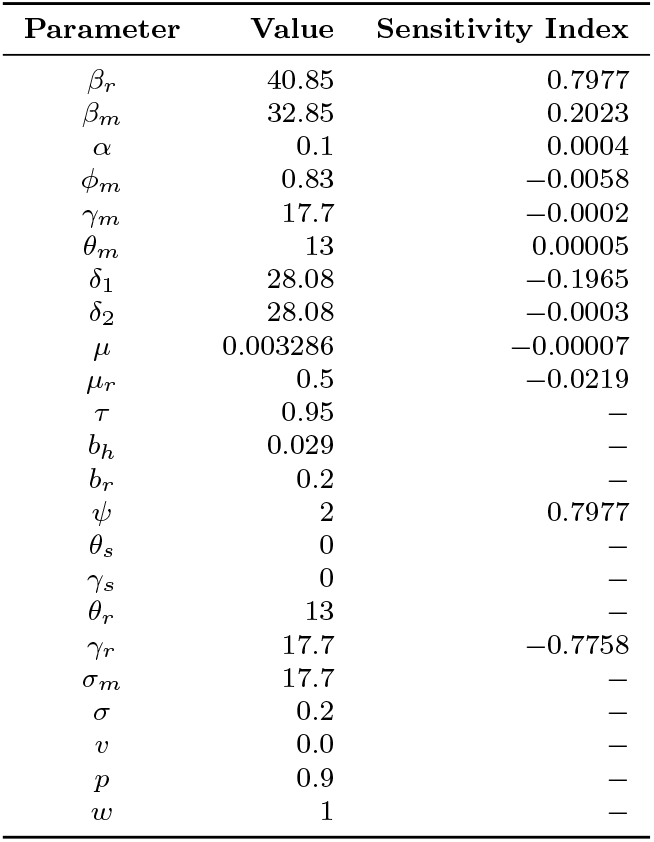
Parameters, Values, and Sensitivity Indices for *R*_0_. The table includes base parameter values and their corresponding forward sensitivity indices computed using Equation (35) in sub-section 4.1

| Parameter | Value | Sensitivity Index |
| --- | --- | --- |
| $\beta_r$ | 40.85 | 0.7977 |
| $\beta_m$ | 32.85 | 0.2023 |
| $\alpha$ | 0.1 | 0.0004 |
| $\phi_m$ | 0.83 | -0.0058 |
| $\gamma_m$ | 17.7 | -0.0002 |
| $\theta_m$ | 13 | 0.00005 |
| $\delta_1$ | 28.08 | -0.1965 |
| $\delta_2$ | 28.08 | -0.0003 |
| $\mu$ | 0.003286 | -0.00007 |
| $\mu_r$ | 0.5 | -0.0219 |
| $\tau$ | 0.95 | — |
| $b_h$ | 0.029 | — |
| $b_r$ | 0.2 | — |
| $\psi$ | 2 | 0.7977 |
| $\theta_s$ | 0 | — |
| $\gamma_s$ | 0 | — |
| $\theta_r$ | 13 | — |
| $\gamma_r$ | 17.7 | -0.7758 |
| $\sigma_m$ | 17.7 | — |
| $\sigma$ | 0.2 | — |
| $v$ | 0.0 | — |
| $p$ | 0.9 | — |
| $w$ | 1 | — |

**Fig. 4:**
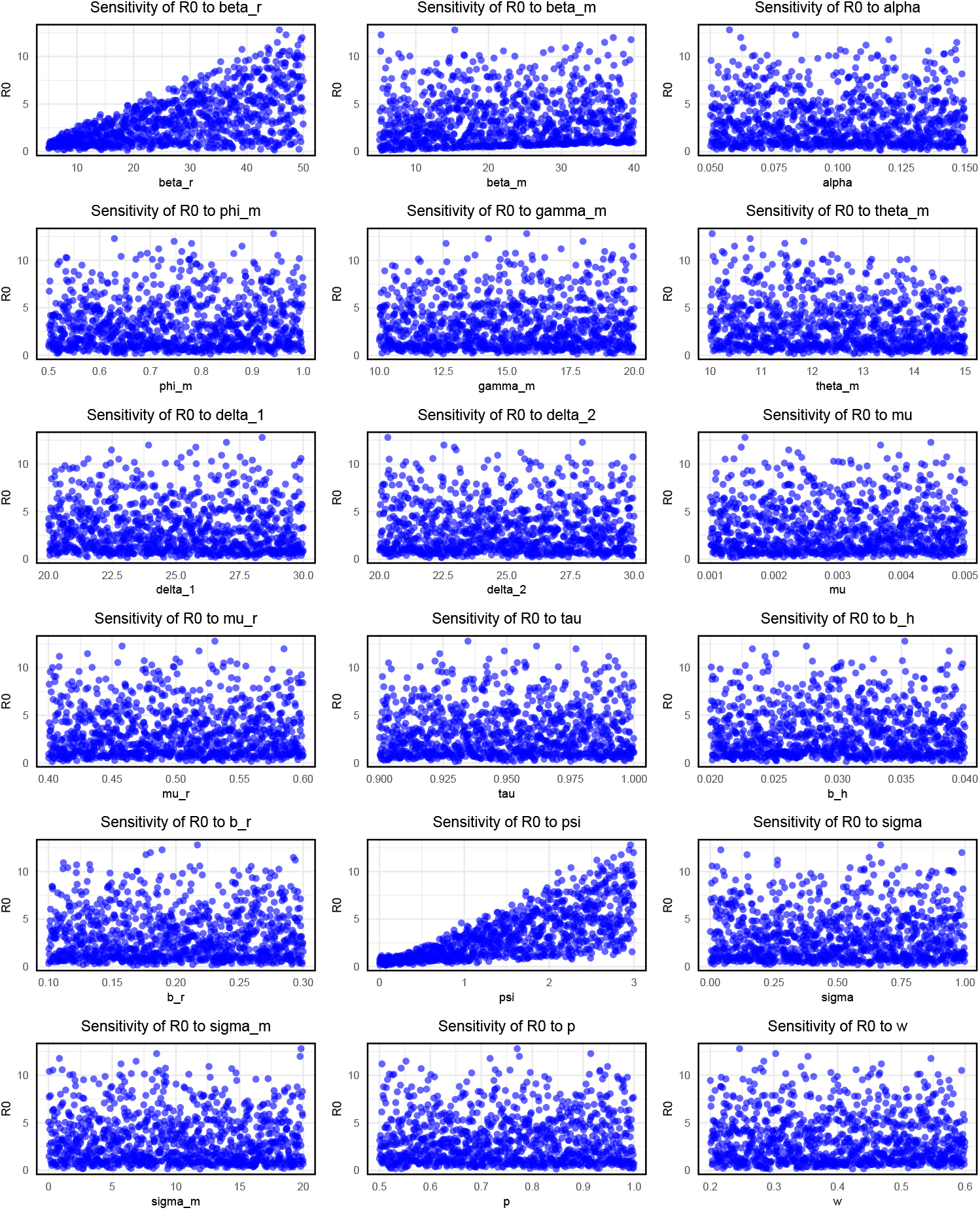
Global Sensitivity Indices for *R*_0_ calculated using Latin Hypercube Sampling (LHS). The indices were computed based on Equations (36) and (37) in Sub-section 4.2, highlighting the relative influence of key parameters on *R*_0_.

The transmission rates of rodents to humans *β*_*r*_ have sensitivity index 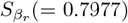. Its high and positive sensitivity index suggests that changes in the rodent to human transmission plays a critical role in the disease dynamics. if *β*_*r*_ increases, the potential for disease spread increases significantly. The sensitivity index for human to human transmission 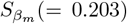 is lower than *β*_*r*_ but still contribute significantly to the spread of monkeypox virus. This suggests that human to human transmission is important, but the rodent to human pathways has stronger influence on the overall transmission dyanamics. This is clearly evident in Figure 4 the increase in the value of *R*_0_ when *β*_*r*_ is increases. The sensitivity index for Relative infectivity rate of rodent to human transmission (*ψ*) is *S*_*ψ*_(= 0.7977). This scaling factor for rodent transmission has a high sensitivity, indicating that the relative infectivity rate of rodent to human transmission is a key factor. This model suggests that control interventions targeting rodents or reducing their population could significantly impact the spread of Monkeypox virus.

Parameters with lower sensitivities includes *α*(*S*_*α*_ = 0.0004), the reduced infectivity rate of hospitalized infectious humans, while recovery rate of hospitalized *γ*_*m*_, has low but negative impact on the dynamics with sensitivity index of *S*_*ψ*_*m* (= −0.0002). The rodent recovery rate of the infected rodents *γ*_*r*_ has a negative sensitivity index *S*_*γ*_*r* (= − 0.7758) which suggests that increasing recovery rate of the infected rodents alters the transmission dynamics and growth of the infection in the human and rodent population. In conclusion, the sensitivity analysis indicates that rodent to human transmission (*β*_*r*_) and the relative infectivity of rodent to human transmission (*ψ*) have the highest impact on *R*_0_. This means that these parameters strongly derive the spread of monkeypox virus. Human-to-Human transmission rate (*β*_*m*_) also plays a significant role but to a lesser extent than rodent to human transmission.

In conclusion, the sensitivity analysis highlights that the parameters *β*_*r*_ and *ψ* are the most significant drivers of Monkeypox virus transmission, with *β*_*m*_ also contributing meaningfully but to a lesser extent. Control measures focusing on reducing rodent-to-human transmission could have the most pronounced impact on controlling the disease.

It is essential to note that the waning immunity from the smallpox vaccine (*σ*) and the loss of temporary immunity from Monkeypox infection (*σ*_*m*_) indirectly affect the susceptible population. Further metrics will be developed to capture their influence on disease dynamics comprehensively.

The following plots (Fig. 5) illustrate the dynamics of the rodent population in a model of monkeypox transmission, with varying transmission rates (*β*_*r*_ = 10, 15, and 20). The plots track the evolution of four key compartments—susceptible (*S*_*r*_), exposed (*E*_*r*_), infectious (*I*_*r*_), and recovered (*R*_*r*_) rodents—over time. The model assumes initial conditions of *S*_*r*_(0) = 5000, *I*_*r*_(0) = 1, *E*_*r*_(0) = 0, and *R*_*r*_(0) = 0. These profiles provide insights into how different transmission rates influence the spread and eventual recovery of the rodent population in the context of the disease dynamics.

**Fig. 5:**
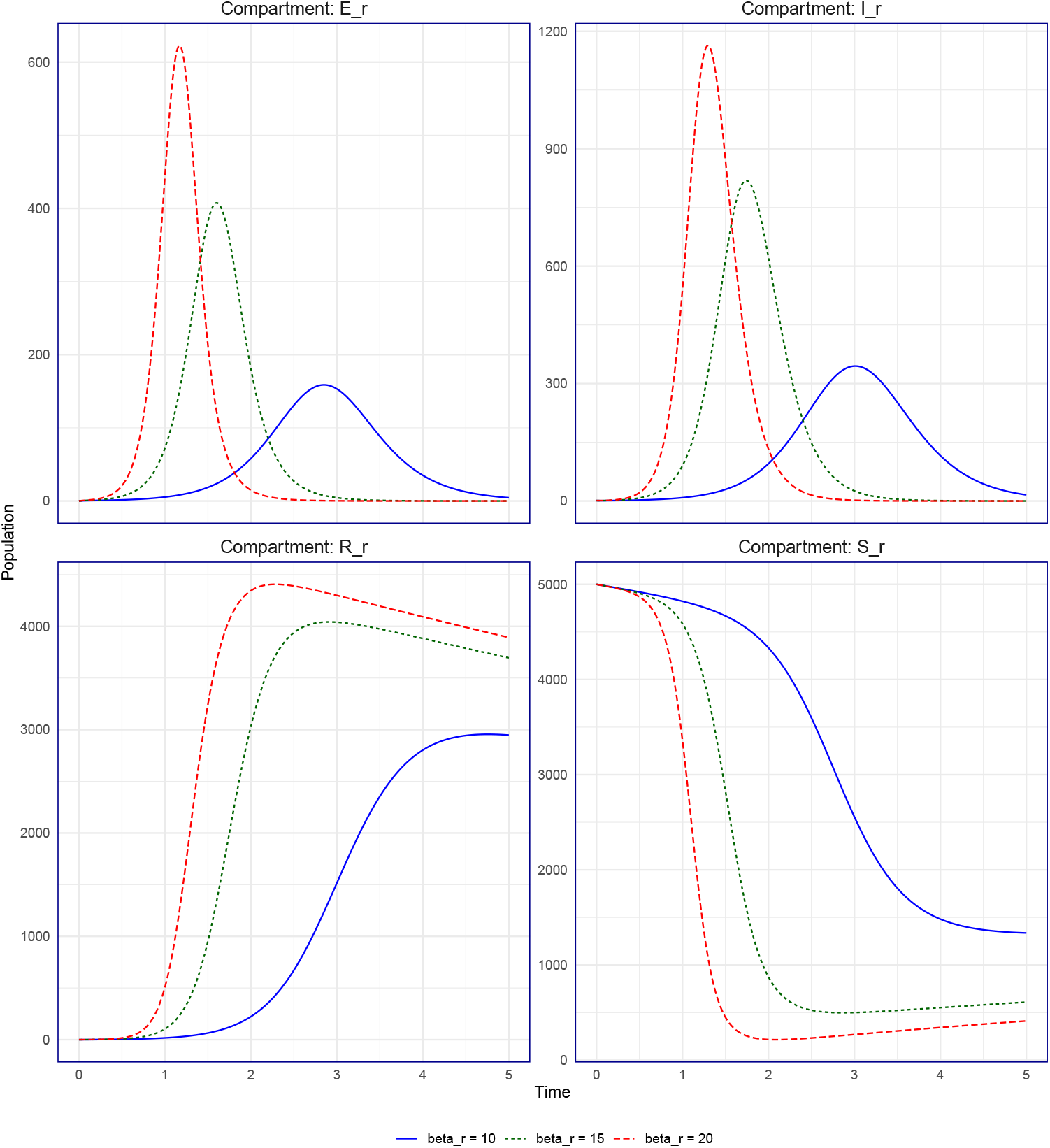
Dynamics of rodent populations under varying transmission rates (*β*_*r*_ = 10, 15, and 20). The profiles illustrate the changes in susceptible (*S*_*r*_), exposed (*E*_*r*_), infectious (*I*_*r*_), and recovered (*R*_*r*_) rodents over time. Initial conditions: *S*_*r*_(0) = 5000, *I*_*r*_(0) = 1, *E*_*r*_(0) = 0, *R*_*r*_(0) = 0.

### 4.3 Threshold theory

The theory of epidemic threshold was first introduced by Kermack and McKendrick (1927). The theory provides a mathematical structure to understand the conditions under which an infectious disease can invade and persist in a population for a general epidemic model (Bailey, 1975). These classical threshold-based models rely on *R*_0_ = 1 as the defining boundary between epidemic and non-epidemic states. Although this threshold is essential, it is insufficient to fully capture the dynamic interplay of immunity, susceptible replenishment, and transmission variability.

Here, we propose a dual threshold theory, as extension of Kermack and McKendrick (1927) that integrates *R*_0_(*t*), the time-dependent reproduction number, and the temporal density of susceptibles *S*_*m*_(*t*), to better characterize epidemic transitions. This formalism is applicable to other diseases, but is particularly applicable to the Monkeypox Virus (MPXV), where waning immunity from vaccine or loss of temporary immunity from disease, zoonotic reservoirs, and demographic replenishment interact to shape outbreak dynamics.

### 4.4 Threshold Densities, Timing and Dynamics of *R*_0_(*t*)

In this study, we examine the time evolution of the basic reproductive number, *R*_0_(*t*), to investigate how variations in population immunity and the waning of vaccine-induced protection influence epidemic dynamics. The analysis establishes a general framework aligned with monkeypox virus (MPXV) transmission dynamics, which can also be broadly applied to other infectious diseases.

The time-dependent reproductive number, *R*_0_(*t*), represents the number of new cases generated by an average case at time *t*, where *t* is measured in units of time (An et al., 2022; Nash et al., 2022). This metric is directly proportional to the density of susceptible individuals at time *t* and is mathematically expressed as:

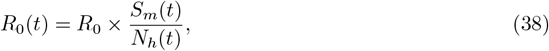

where *R*_0_ is the basic reproduction number in a fully susceptible population, *S*_*m*_(*t*) denotes the density of susceptibles, and *N*_*h*_(*t*) represents the total host population at time *t*. The ratio *R*_0_*/N*_*h*_(*t*) is the scaling factor.

Equation (34) unifies *R*_0_(*t*), *R*_0_, and *S*_*m*_(*t*), emphasizing the interplay between the time-dependent reproduction number, the basic reproduction number, and the dynamics of the susceptible population.

#### Temporal Condition for Epidemic Growth

The temporal evolution of the basic reproductive number, *R*_0_(*t*), is fundamental to understanding the progression of an epidemic. A critical determinant in this process is the initial growth rate of *R*_0_(*t*). For *R*_0_(*t*) to increase, the condition 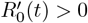 must hold, where:

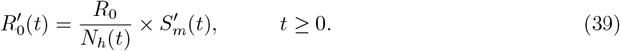

From Equation (39), the inequality 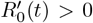 implies that the time-dependent reproductive number *R*_0_(*t*) increases when the susceptible population exhibits a positive growth rate 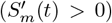. This emphasizes the direct relationship between the growth of *R*_0_(*t*) and the increasing density of susceptible individuals. This relationship underscores the critical role of the dynamics of *S*_*m*_(*t*) in influencing changes in *R*_0_(*t*), thereby highlighting its importance in shaping epidemic trajectories.

### 4.5 Dual Threshold Theory

The Dual threshold theory, provides significant conceptual advancement in the Kermack and McK-endrick (1927) threshold theory by incorporating two independent thresholds, the temporal basic reproductive number *R*_0_(*t*) and the density of susceptibles *S*_*m*_(*t*).

A comprehensive analysis is presented using Figure 6, which depicts the time-dependent reproductive number *R*_0_(*t*) as a function of time *t*. This plot is critical for understanding the current epidemic state, as it characterizes the behavior of the solution curves for *R*_0_(*t*). The graph delineates two distinct regions separated by the threshold line at *R*_0_(*t*) = 1. The upper region, referred to as the “epidemic region,” represents conditions where *R*_0_(*t*) > 1, indicating a high risk of sustained epidemic spread. In contrast, the lower region, the “non-epidemic region,” corresponds to *R*_0_(*t*) < 1, where the transmission rate is insufficient to sustain an outbreak, and the epidemic risk is minimal. The threshold line *R*_0_(*t*) = 1 marks a critical transition between these regions. It signifies the point where the system stabilizes and potentially reaches endemic equilibrium. When *R*_0_(*t*) > 1, the reproductive number exceeds the critical value necessary for disease propagation, elevating the risk of an epidemic. Conversely, when *R*_0_(*t*) < 1, the transmission dynamics favor epidemic decline, and the outbreak tends to subside. The basic reproductive number *R*_0_(*t*) plays a central role in governing the dynamics of an epidemic. For an epidemic to occur, *R*_0_(*t*) > 1 at any given time *t*, satisfying the inequality:

**Fig. 6:**
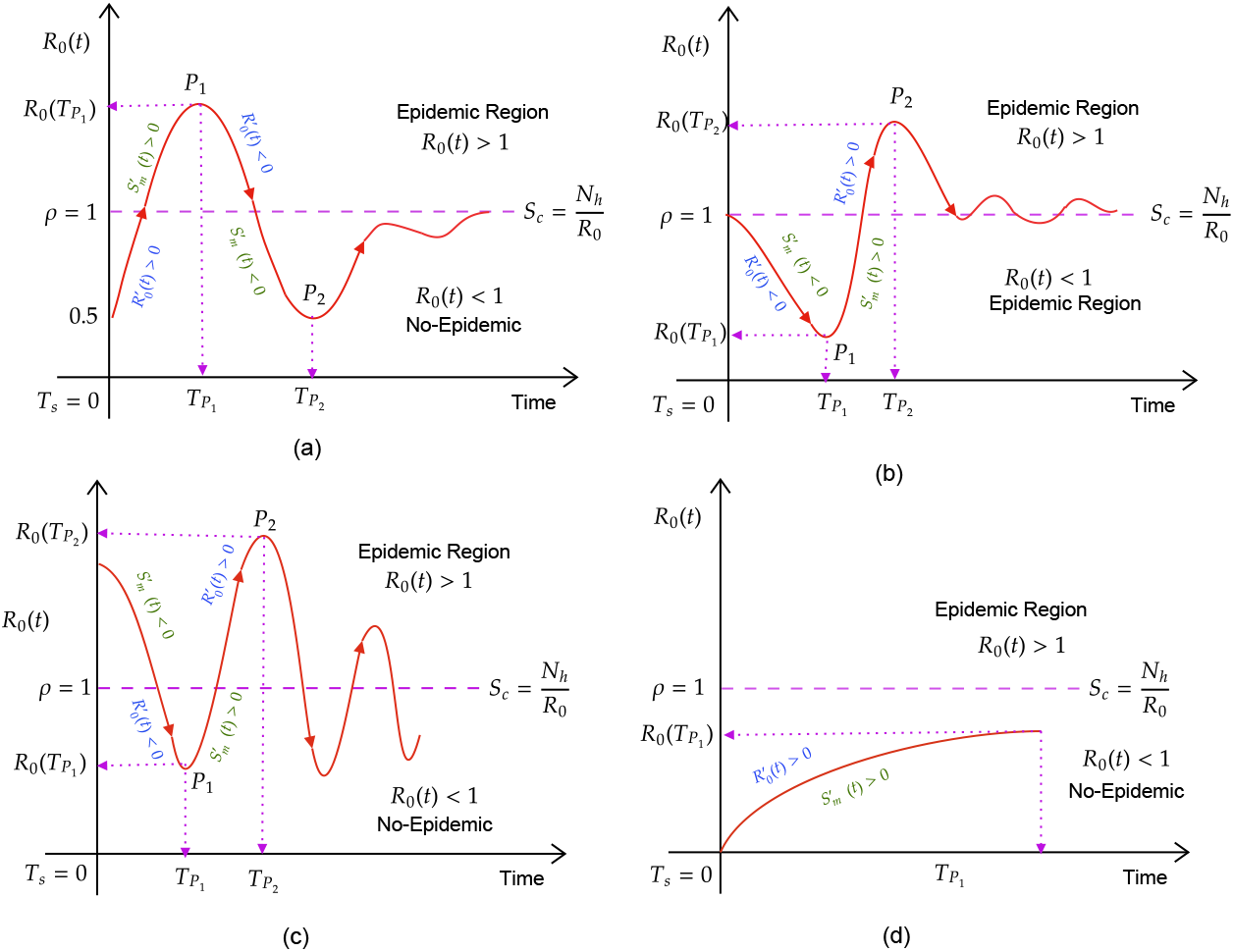
Time-dependent reproductive number *R*_0_(*t*) and susceptible population *S*_*m*_(*t*) with associated thresholds and regions. The figure delineates the epidemic (*R*_0_(*t*) > 1 and *S*_*m*_(*t*) > *S*_*c*_(*t*)) and non-epidemic (*R*_0_(*t*) ≤ 1 or *S*_*m*_(*t*) ≤ *S*_*c*_(*t*)) regions, highlighting the critical thresholds that govern the transition between these regions. The threshold lines (*R*_0_(*t*) = 1 and *S*_*m*_(*t*) = *S*_*c*_(*t*)) are key to understanding the conditions necessary for epidemic initiation and progression.

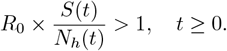

This condition implies that the density of susceptibles must exceed the critical threshold, *S*(*t*) > *N*_*h*_(*t*)*/R*_0_. When *S*(*t*) surpasses this threshold, the system transitions into the epidemic region, characterized by *R*_0_(*t*) > 1. The system remains in this region until the force of infection decreases or the susceptible population becomes sufficiently depleted. Conversely, the system remains in the non-epidemic region when *R*_0_(*t*) < 1, satisfying the inequality:

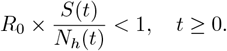

This condition is met when *S*(*t*) < *N*_*h*_(*t*)*/R*_0_. If the system initially resides in the non-epidemic region, it will remain there as long as *S*(*t*) remains below this threshold. This theory provides valuable insights into the current epidemic state, determining whether a disease will persist (*R*_0_(*t*) > 1) or fade out (*R*_0_(*t*) < 1). Threshold theory is instrumental in identifying critical parameters, such as vaccination coverage or booster frequency, needed to maintain *R*_0_(*t*) < 1 and ensure long-term epidemic control.

The dual threshold aligns with the fundamental principles of infectious disease dynamics in classical epidemic theory. We will highlight the importance of the dual threshold theory and the epidemiological implications as follows:

1. **Threshold 1:** *R*_0_(*t*) = 1 **and** *S*(*t*) = *N*_*h*_(*t*)*/R*_0_: This threshold gives the boundry between epidemic and non-epidemic regions. This gives the minimum condition for an infectious disease to spread in a population.
  i. This theory recognizes that *R*_0_(*t*) = 1, the classic threshold for sustained transmission is necessary but not sufficient to trigger or sustain epidemics.
  ii. By introducing the second threshold, *S*(*t*) = *N*_*h*_(*t*)*/R*_0_, addresses the critical density of susceptible required for epidemic growth.
2. **Threshold 2:** *R*_0_(*t*) > 1 **and** *S*(*t*) ≤ *N*_*h*_(*t*)*/R*_0_: The threshold number of susceptible are not sufficient to withstand an epidemic, hence there will be low transmission with minor epidemic and when the number of susceptible is sufficiently low, and *R*_0_(*t*) < 1 the system will remain in the non-epidemic region.
3. **Threshold 3:** *R*_0_(*t*) > 1 **and** *S*(*t*) ≤ *N*_*h*_(*t*)*/R*_0_: Since the the time-dependent basic reproductive is greater one (*R*_0_(*t*) > 1) at any time t, and the threshold density of susceptible is greater than the the ratio of total population density and static reproductive number *R*_0_, the system will starts its dynamics in the epidemic region.

It is important to note the relationship between *R*_0_(*t*) and *S*(*t*). the first is that the condition *R*_0_(*t*) > 1 is necessary but not sufficient for an epidemic. It must be coupled with *S*(*t*) > *N*_*h*_(*t*)*/R*_0_ to ensure sustain transmission, secondly, as *S*(*t*) = *N*_*h*_(*t*)*/R*_0_, the system lies exactly on the threshold, with the disease dynamics being in a marginal state which can lead to minor epidemic.

#### 4.5.1 Temporal Dynamics and Dual Threshold

The dynamics of the dual threshold depend on the initial reproductive number, *R*_0_(*T*_*s*_ = 0), and the initial density of susceptibles, *S*_*m*_(*T*_*s*_ = 0). Here, *T*_*s*_ represents the initial time at which *R*_0_(*t*) and *S*_*m*_(*t*) begin to evolve, either increasing or decreasing, within the epidemic or non-epidemic region, as illustrated in Figure 6. This section explores the properties of these threshold conditions:

##### Case I: *S*_*m*_(*T*_*s*_ = 0) < *S*_*c*_(*T*_*s*_ = 0), *R*_0_(*T*_*s*_ = 0) < 1

As shown in Fig. 6(a), the system starts at *T*_*s*_ in the non-epidemic region, where 0 < *R*_0_(*t*) ≤ 1*/*2. At this stage, *R*_0_(*t*) and *S*_*m*_(*t*) begin to increase 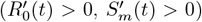.The transition across the threshold line depends on the critical density of susceptibles (*S*_*c*_(*T*_*s*_ = 0) = *N*_*h*_(*T*_*s*_ = 0)*/R*_0_). Once the threshold line (*ρ* = 1) is crossed, the threshold condition is met, and *R*_0_(*t*) reaches a peak at the point *P*_1_, with a corresponding temporal value *R*_0_(*T*_*P*_1), where *T*_*P*_1 denotes the time at which *R*_0_(*t*) and *S*_*m*_(*t*) are maximum simultaneously.

At *P*_1_, the system reaches its maximum within the epidemic region, with *R*_0_(*t*) and *S*_*m*_(*t*) attaining their highest values. At this point, the derivatives satisfy 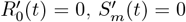,while the second derivatives indicate a concave-down behavior 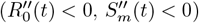.

Since the density of susceptibles at *P*_1_ is sufficiently high (greater than the threshold *S*_*c*_(*t*)), the system initiates another cycle, eventually descending to the point *P*_2_ in the non-epidemic region. At this stage, the system decelerates, with 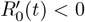 and 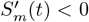,and reaches the turning point *P*_2_ at time 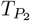.

At *P*_2_, *R*_0_(*t*) and *S*_*m*_(*t*) attain their minimum values, with 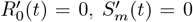, and second derivatives indicating a concave-up behavior 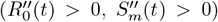. Notably, this point corresponds to the maximum number of infectives, *I*_*m*_(*t*). Beyond *P*_2_, the density of susceptibles becomes insufficient (*S*_*m*_(*t*) < *S*_*c*_(*t*)), and the system remains within the non-epidemic region.

##### Case II: *S*_*m*_(*T*_*s*_ = 0) ≈ *S*_*c*_(*T*_*s*_ = 0), 0 < *R*_0_(*T*_*s*_ = 0) ≤ 1

The threshold condition for this scenario is illustrated in Figure Fig. 6(b). At *T*_*s*_ = 0, the reproductive number is at the threshold line, *R*_0_(*T*_*s*_) = *ρ* = 1, and the initial density of susceptibles, *S*_*m*_(*T*_*s*_ = 0), is approximately equal to the critical threshold, *S*_*c*_ = *N*_*h*_*/R*_0_.

In this case, the density of susceptibles is large but not sufficient to trigger a major epidemic. Instead, a minor epidemic is initiated as *R*_0_(*t*) begins to decrease 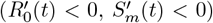,starting from *T*_*s*_ = 0.

The system reaches its first turning point, *P*_1_, at time *T*_*P*_1 within the non-epidemic region. At *P*_1_, the susceptibles are nearly exhausted, and the reproductive number is at its minimum. The second derivatives at this point indicate a concave-up behavior 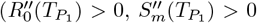.Furthermore, this point corresponds to the maximum number of infectives (*I*_*m*_(*t*)), where the second derivative of infectives is negative 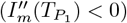.

##### Case III: *S*_*m*_(*T*_*s*_ = 0) > *S*_*c*_(*T*_*s*_ = 0), *R*_0_(*T*_*s*_ = 0) > 1

Fig. 6 (c) illustrates a scenario that begins in the epidemic region, where the susceptible population is sufficiently high to initiate an epidemic, as it exceeds the critical threshold density of susceptibles (*S*_*m*_(*T*_*s*_) > *N*_*h*_(*T*_*s*_), *R*_0_(*T*_*s*_) > 1). The epidemic starts at *T*_*s*_ = 0, when both the reproductive number and the susceptible population begin to decline 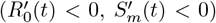.Over time, this depletion drives the system into the non-epidemic region, reaching point *P*_1_ at time 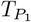.

Fig. 6(c) also exhibits multiple cycles, which occur due to the replenishment of the susceptible population. As the system transitions back toward the epidemic region, the number of susceptibles and the reproductive number increase over time. However, once the replenishment of susceptibles ceases (*S*_*m*_(*t*) < *N*_*h*_(*t*), *R*_0_(*t*) < 1) for all time *t*, the disease eventually dies out.

##### Case IV: *S*_*m*_(*T*_*s*_ = 0) < *S*_*c*_(*T*_*s*_ = 0), *R*_0_(*T*_*s*_ = 0) < 1

Fig. 6(d) illustrates a scenario where no epidemic is initiated, as *R*_0_(*T*_*s*_) remains in the non-epidemic region throughout the entire period. This occurs because the number of susceptibles is insufficient to sustain the epidemic (*S*_*m*_(*T*_*s*_) < *N*_*h*_(*T*_*s*_), *R*_0_(*T*_*s*_) < 1). Once the few individuals in the susceptible compartment become infected, the disease dies out.

### 4.6 Threshold Condition for MPXV Model

The threshold condition derived in section 4.1-4.3 is central to understanding the dynamics of Monkeypox virus (MPXV) transmission over time. It provides a mathematical framework for evaluating the temporal impact of waning immunity on the risk of MPXV epidemics. Specifically, this condition helps determine whether the Monkeypox virus will persist and spread within a population or eventually fade out. By analyzing the threshold over time, the model captures how shifts in immunity levels influence outbreak potential and shape the broader transmission dynamics.

#### 4.6.1 Starting in the Epidemic Region: *S*_*m*_(*T*_*s*_) > *S*_*c*_(*T*_*s*_), *R*_0_(*T*_*s*_) > 1

Starting at the initial time *T*_*s*_ = 0, an epidemic is initiated when *R*_0_(*T*_*s*_ = 0) is in the epidemic region. This occurs when the number of susceptibles at the starting time, *S*_*m*_(*T*_*s*_ = 0), is above the threshold density of susceptibles (*S*_*c*_(*T*_*s*_ = 0) = *N*_*h*_(*T*_*s*_ = 0)*/R*_0_), and the initial reproductive number satisfies *R*_0_(*T*_*s*_ = 0) > 1. This corresponds to scenario III, where an epidemic is immediately triggered due to these conditions.

When the epidemic begins, the number of susceptibles (*S*_*m*_(*t*)) and the time-dependent reproductive number (*R*_0_(*t*)) start to decline due to the pressure of infection 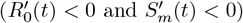. Simultaneously, the number of infectives (*I*_*m*_(*t*)) begins to increase from the initial time point *T*_*s*_ = 0. The derivative of the time-dependent reproductive number is given by:

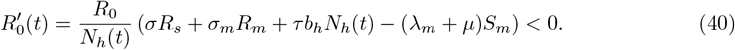

From this inequality, we have the following condition:

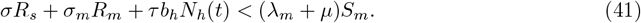

This inequality provides the condition under which the number of susceptibles and the time-dependent reproductive number decrease after the epidemic is initiated. The condition for an epidemic to initiate requires the number of infectives to grow 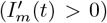.This condition is satisfied when the rate of transition from the exposed to the infectious compartment exceeds the combined rate of recovery, death, and progression to other compartments for infective individuals. Mathematically, this is expressed as:

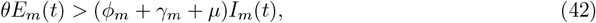

where *θ* represents the rate of progression from the exposed to infectious class, *E*_*m*_(*t*) is the number of exposed individuals at time *t, ϕ*_*m*_ is the rate of progression to severe cases, *γ*_*m*_ is the recovery rate, and *μ* is the natural mortality rate. This inequality highlights the critical balance required for the infectious population to increase, thereby initiating an epidemic.

Here, the growth of the infective population occurs when the number of individuals transitioning from the exposed compartment (*E*_*m*_) to the infectious compartment exceeds the number of individuals leaving the infectious compartment (*I*_*m*_) at any time *t*. The condition for starting an epidemic can also be described in terms of a threshold ratio:

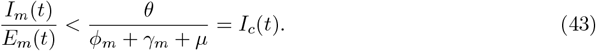

This indicates that the ratio of infectious individuals to exposed individuals at time *t* must be less than the threshold *I*_*c*_(*t*), which represents the number of infectious individuals transitioning from the exposed class over the average duration of infectiousness.

In conclusion, an epidemic is initiated when the following conditions are satisfied:

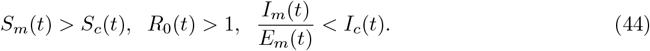

These results allow us to deduce the following:

i. When *R*_0_(*t*) starts in the epidemic region (*S*_*m*_(*T*_*s*_ = 0) > *S*_*c*_(*T*_*s*_ = 0), *R*_0_(*T*_*s*_ = 0) > 1), the epidemic is immediately initiated. The force of infection acts directly on the susceptibles, causing a decline in their number, *S*_*m*_(*t*), and the time-dependent reproductive number, *R*_0_(*t*). This results in the density of susceptibles falling below the threshold density, *S*_*c*_(*t*).
ii. In Equation (**??**) the condition for decrease in the number of susceptibles and *R*_0_(*t*) is satisfied when the replenishment of susceptibles, either through waning immunity or natural birth, is less than the losses due to natural death or infection.
iii. The minimum values of *R*_0_(*t*) and *S*_*m*_(*t*) represent the turning points in the non-epidemic region. At the minimum point described in (iii), *I*_*m*_(*t*) reaches its maximum. The time point *T*_*p*_, at which *I*_*m*_(*t*) peaks, is defined as the peak time of the epidemic. At this point:

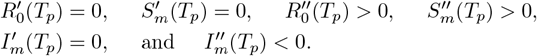

For the first case 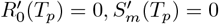,we have

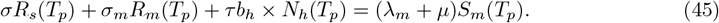

The condition in Equations (41) and (45) are satisfied for both *S*_*m*_(*t*) and *R*_0_(*t*). It represents the equality between the replenishment of susceptibles through waning immunity or natural birth and the loss of susceptibles through infection or natural death.

The second condition, 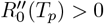 and 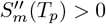 can be expressed as:

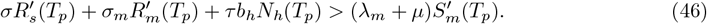

Substituting the corresponding differential equations into Equation (46) and simplifying, we obtain:

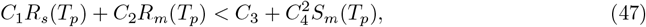

where the coefficients are defined as follows:

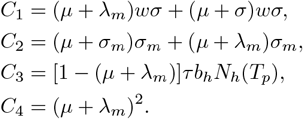

The epidemiological implication of Equation (47) is that the number of individuals in the immune class at the peak time of the epidemic is less than the minimum number of susceptibles at the same peak time, *T*_*p*_. This indicates there is no replenishment of susceptibles at that time.

The conditions 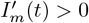 and 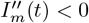 at time *t* = *T*_*p*_ can be expressed as:

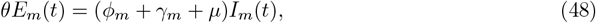

and

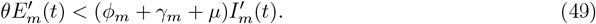

Equation (48) is straightforward; it is satisfied when *I*_*m*_(*t*)*/E*_*m*_(*t*) < *I*_*c*_(*t*). Equation (49) is obtained by substituting expressions for 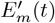 and 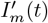,yielding:

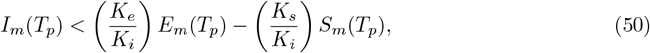

where the coefficients are constants, defined as

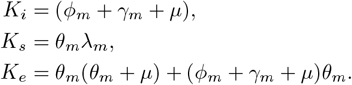

Here:

i. *I*_*m*_(*T*_*p*_): The number of cases infected at peak time *T*_*p*_.
ii. (*K*_*e*_*/K*_*i*_)*E*_*m*_(*T*_*p*_): The number of exposed individuals at the peak time.
iii. (*K*_*s*_*/K*_*i*_)*S*_*m*_(*T*_*p*_): The number of new infections at the peak time *T*_*p*_.

#### 4.6.2 Starting in the Non-Epidemic Region: *S*_*m*_(*T*_*s*_) < *S*_*c*_(*T*_*s*_), *R*_0_(*T*_*s*_) < 1

When the initial density of susceptibles (*S*_*m*_(*T*_*s*_ = 0)) is less than the critical density of susceptibles (*S*_*c*_ = *N*_*h*_(*T*_*s*_ = 0)*/R*_0_) required to start an epidemic, and the initial time-dependent reproductive number (*R*_0_(*T*_*s*_ = 0)) is below the critical threshold 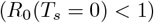, the population lacks sufficient susceptibles to initiate an epidemic. However, as time progresses, both *R*_0_(*t*) and *S*_*m*_(*t*) begin to increase (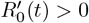and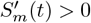) due to processes such as natural births or waning immunity. This growth drives the system toward the epidemic region. Once the threshold conditions (*R*_0_(*t*) = 1 and *S*_*m*_(*t*) = *S*_*c*_(*t*)) are met, the system crosses into the epidemic region. At this point, an epidemic can be initiated at any time when these conditions are satisfied.

The functional relationship between *S*_*m*_(*t*) and *R*_0_(*t*) allows us to express the growth of *R*_0_(*t*) and *S*_*m*_(*t*) as:

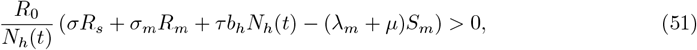

where

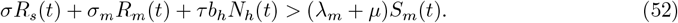

The inequality in Equation (52) provides the threshold condition for the growth of the susceptible population (*S*_*m*_(*t*)) and the time-dependent reproductive number (*R*_0_(*t*)) that will lead the system into the epidemic region.

Upon crossing the epidemic threshold (*S*_*m*_(*t*) > *S*_*c*_(*t*) and *R*_0_(*t*) > 1), and as the susceptible population and the effective reproductive number peak and begin to decline, an epidemic can be initiated when 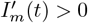,that is:

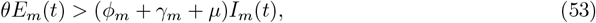

and reaches its maximum when:

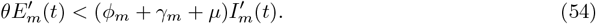

This condition is equivalent to Equations (44, 45, and 46), and the same conclusions can be drawn:

1. Epidemic growth is driven by the transition of exposed individuals (*E*_*m*_(*t*)) into the infectious class, with the condition for growth being 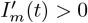.
2. The epidemic reaches its peak when 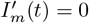 and 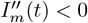.
3. The replenishment of susceptibles through waning immunity or births sustains the system until it crosses into the epidemic region.
4. The smaller the initial reproductive number, the higher the threshold density of susceptible is needed to crosses the threshold boundary for epidemic to be triggered.

The inequality in Equation (44) serves as a critical condition for the system to approach the epidemic region. The epidemic starts only after *R*_0_(*t*) > 1 and *S*_*m*_(*t*) > *S*_*c*_(*t*), while infectious growth conditions 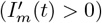 further sustain the outbreak and epidemic peaks when 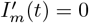, which aligns with the minimum of *S*_*m*_(*t*) and *R*_0_(*t*), as established in earlier results.

### 4.7 Dynamics of Epidemic Thresholds on compartmental profiles

Understanding the dynamics of infectious diseases is critical for predicting outbreaks and developing effective control strategies. The basic reproductive number, *R*_0_(*t*), plays a fundamental role in determining the potential for an epidemic to occur. Specifically, when *R*_0_(*t*) > 1, an epidemic can sustain itself, while *R*_0_(*t*) < 1 leads to the eventual elimination of the infection. Threshold theory provides a valuable framework for analyzing how the interaction of epidemiological parameters, including waning immunity and population susceptibility, influences these dynamics.

This study examines how threshold theory influences the dynamics of *R*_0_(*t*), *S*_*m*_(*t*) (susceptible individuals), *R*_*s*_(*t*) (immune individuals due to smallpox vaccination), and *I*_*m*_(*t*) (infectious individuals). The critical threshold is reached when *R*_0_(*t*) crosses 1, a shift driven by changes in the susceptible population (*S*_*m*_(*t*)) relative to the critical susceptible threshold (*S*_*c*_(*t*)). Understanding the interactions between these variables is crucial for exploring how epidemics progress and the conditions that allow outbreaks to either persist or be controlled.

Fig. 7-9 illustrates three key scenarios with varying levels of population transmission, represented by *R*_0_ and *ψ*. Fig. 7 begins with a low transmission scenario (*R*_0_ < 1 for *ψ* = 0), where transmission occurs solely among humans, without rodent interaction. The curves are plotted for three initial values of susceptibles (*S*_*m*_(0) = 0, 2500, and10000), while the initial number of individuals immune to MPXV due to smallpox vaccination (*R*_*s*_(0) = 10000) remains constant across all plots. Fig. 8 represents a moderate transmission scenario (*R*_0_ for *ψ* = 0.5), where infections occur both between humans and from rodents to humans, with the relative infectivity of rodents to humans being half that of human-to-human transmission. The plots use the same initial number of susceptibles (*S*_*m*_(0)) and immune/vaccinated individuals (*R*_*s*_(0)) as in the low transmission scenario. Fig. 9 depicts a high transmission scenario (*R*_0_ for *ψ* = 1), with equal transmission rates between rodents and humans, and human-to-human. All scenarios (low to high transmission) utilize similar initial conditions. These three cases explore varying levels of immunity—both due to smallpox vaccination (*σ*) and recovery from previous infection (*σ*_*m*_)—to highlight the impact of waning immunity on the threshold dynamics of MPXV epidemics.

**Fig. 7:**
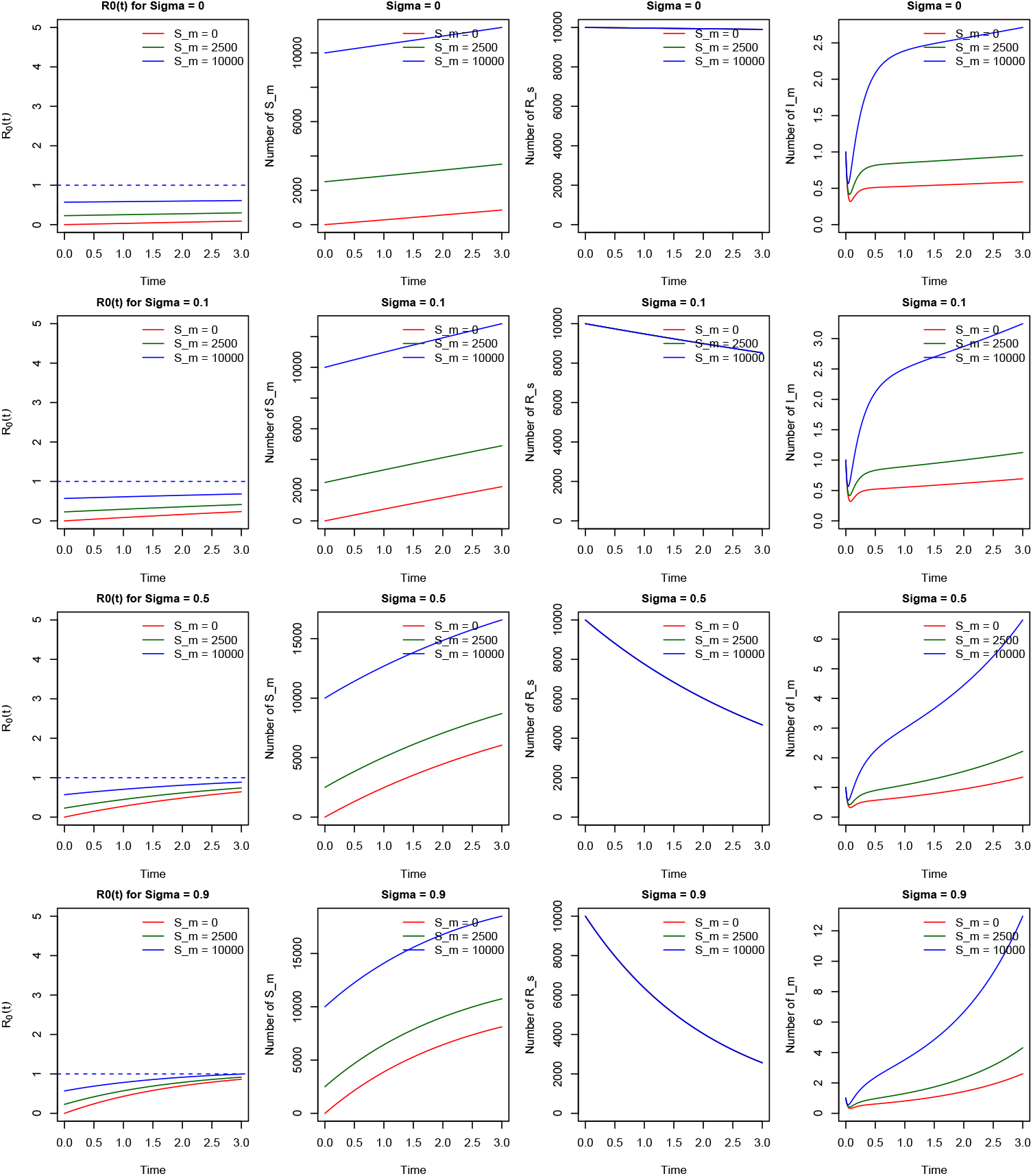
Dynamics of *R*_0_(*t*), *S*_*m*_(*t*), *R*_*s*_(*t*), and *I*_*m*_(*t*) under varying levels of waning immunity (*σ* = 0, 0.1, 0.5, and 0.9) for low transmission potential. The plot illustrates the effect of waning immunity on the threshold dynamics when *R*_0_(0) < 1, with initial conditions *S*_*m*_(0) = 0, 2500, and 10000, *R*_*s*_(0) = 10000, and the relative infectivity rate of rodent to human is *ψ* = 0.

**Fig. 8:**
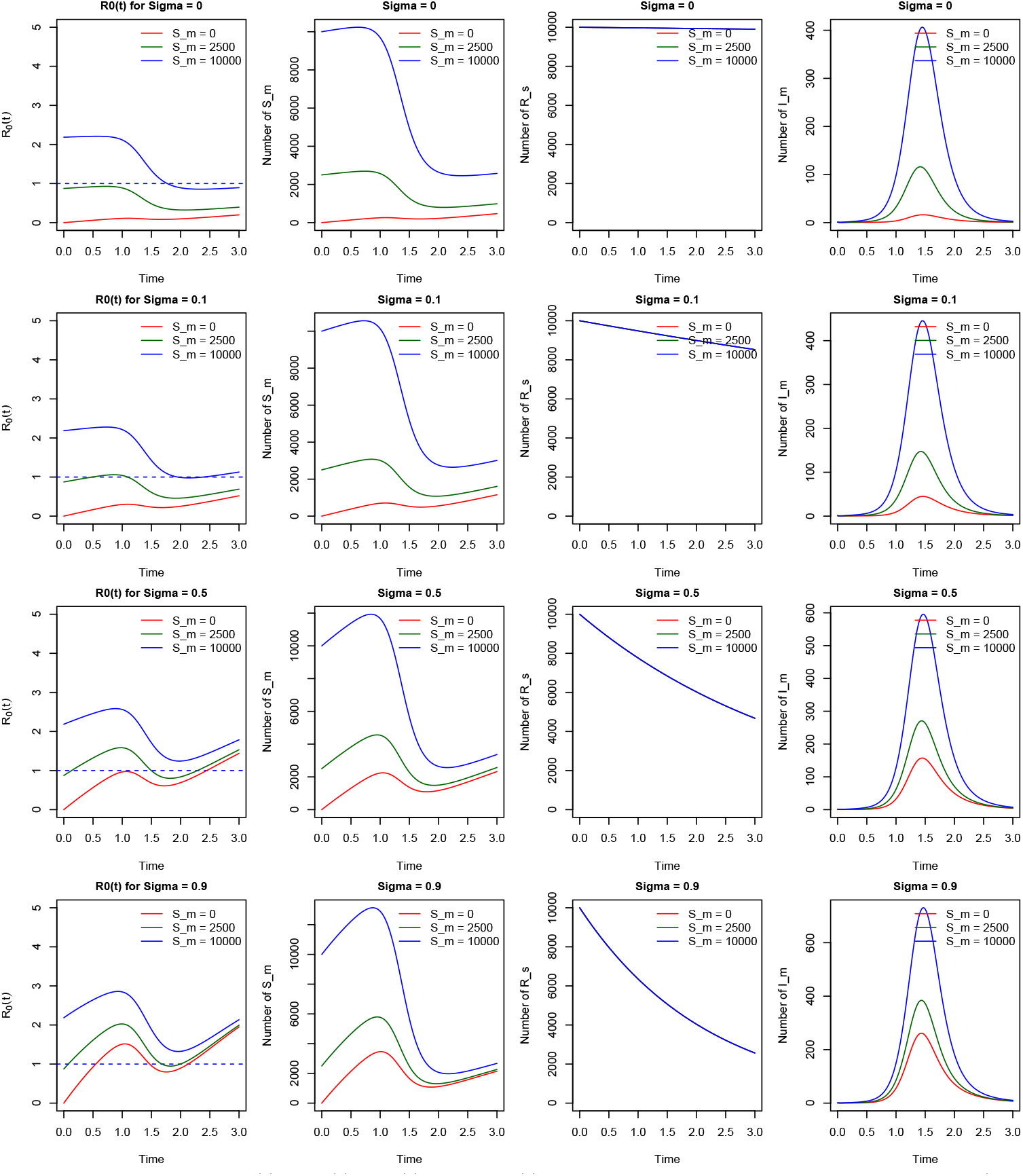
Dynamics of *R*_0_(*t*), *S*_*m*_(*t*), *R*_*s*_(*t*), and *I*_*m*_(*t*) under varying levels of waning immunity (*σ* = 0, 0.1, 0.5, and 0.9) for moderate transmission potential. The plot examines the effect of waning immunity on the threshold dynamics with initial conditions *S*_*m*_(0) = 0, 2500, and 10000 and *R*_*s*_(0) = 10000. For *S*_*m*_(0) = 10000, *R*_0_(0) > 1, whereas for *S*_*m*_(0) = 0, 2500, *R*_0_(0) ≤ 1. The relative infectivity rate of rodent to human is *ψ* = 0.5.

**Fig. 9:**
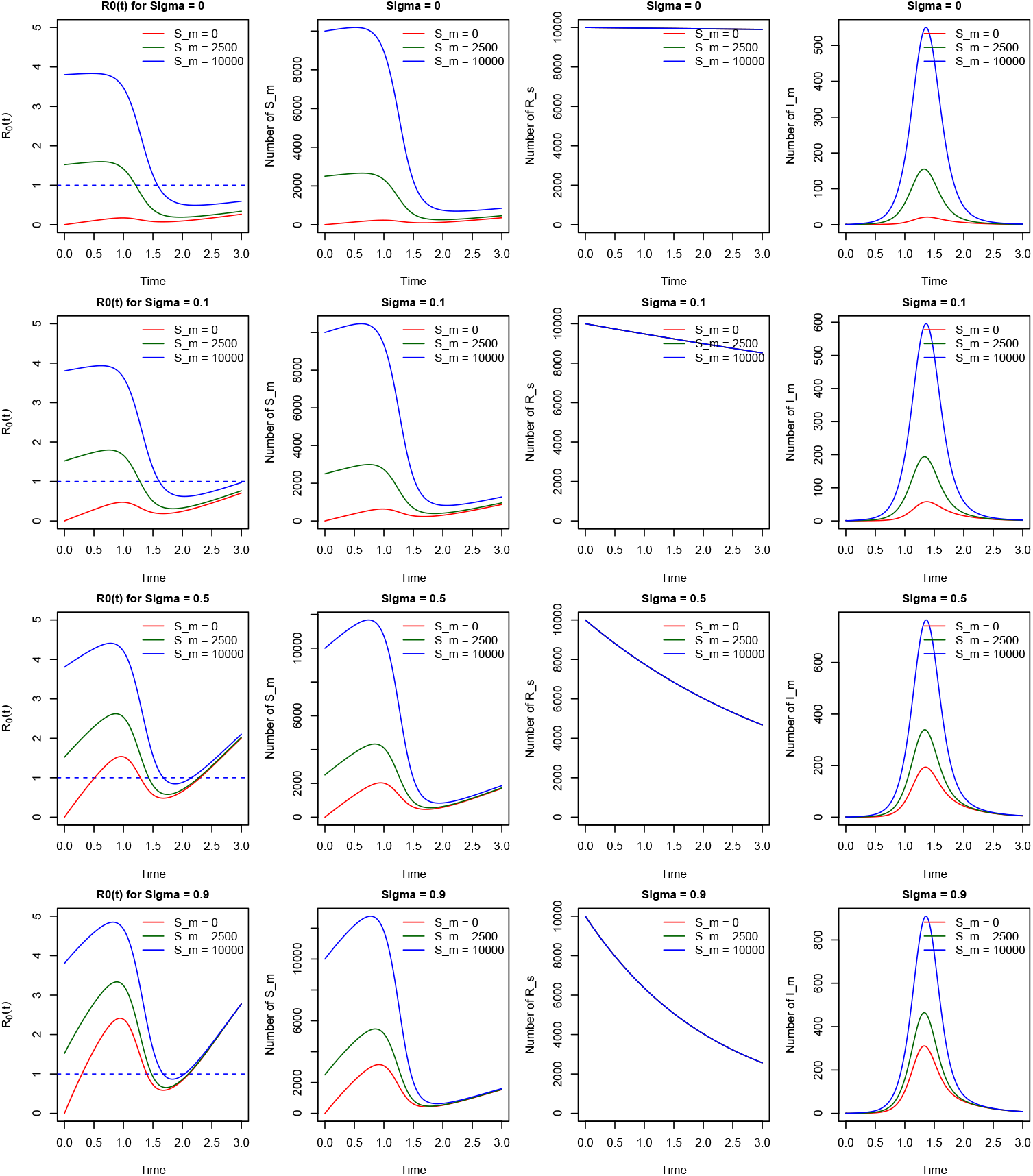
Dynamics of *R*_0_(*t*), *S*_*m*_(*t*), *R*_*s*_(*t*), and *I*_*m*_(*t*) under varying levels of waning immunity (*σ* = 0, 0.1, 0.5, and 0.9) for High transmission potential. The plot examines the effect of waning immunity on the threshold dynamics with initial conditions *S*_*m*_(0) = 0, 2500, and 10000 and *R*_*s*_(0) = 10000. For *S*_*m*_(0) = 2500, 10000, *R*_0_(0) > 1, whereas for *S*_*m*_(0) = 0, *R*_0_(0) < 1.The relative infectivity rate of rodent to human is *ψ* = 1.

In Fig. 7, examining the dynamics of *R*_0_(*t*) and the infective profile *I*_*m*_(*t*) over time reveals that no epidemic is initiated for any of the three initial densities of susceptibles. All curves remain within the non-epidemic region throughout the given period. Even with a high initial density of susceptibles, an epidemic fails to occur. This phenomenon arises because the small initial value of *R*_0_ < 1 results in a critical threshold density of susceptibles that is too large to be reached, preventing the epidemic threshold from being crossed. Here, despite the impact of waning immunity on the dynamics of the susceptible population, neither permanent immunity (*σ* = 0) nor waning immunity (*σ* > 0) is sufficient to trigger an epidemic. This is primarily due to the low transmission rate associated with human-to-human interactions alone. This underscores the critical role of rodents in amplifying the risk of an epidemic, as demonstrated by the sensitivity analysis results.

In Fig. 8, for the scenario with a low initial number of susceptibles (*S*_*m*_(0) = 0), the trajectory of *R*_0_(*t*) under permanent immunity (*σ* = 0) shows that no epidemic is initiated, even with a moderate reproductive number. This is due to the insufficient initial susceptible population, which keeps the system within the non-epidemic region. The infective profile (*I*_*m*_(*t*)) further supports this observation, as no significant rise in infections is observed. Additionally, there is no reduction in the immune population (*R*_*s*_(*t*)) because permanent immunity (*σ* = 0) prevents waning, thereby eliminating any replenishment of the susceptible population from the immune class (*R*_*s*_(*t*)) that could potentially increase *R*_0_(*t*) and push the system across the epidemic threshold.

By increasing the waning rate to *σ* = 0.5 and *σ* = 0.9, for *S*_*m*_(0) = 0, 2500, and 10000, a minor epidemic is initiated. This is due to the higher waning rate, which results in a substantial number of immune individuals losing immunity, thereby increasing the susceptible population and pushing the system across the threshold boundary. The infective profile plots illustrate that, for all levels of *σ*, when *S*_*m*_(0) = 10000, the system begins in the epidemic region. The high initial number of susceptibles leads to a significant epidemic, with the number of cases at the peak increasing as the waning rate increases. Similarly, for *S*_*m*_(0) = 2500, the system crosses the epidemic threshold only for *σ* = 0.5 and *σ* = 0.9, indicating that waning immunity plays a critical role in driving the epidemic dynamics in these scenarios.

Fig. 9 presents the case for high transmission, where equal relative transmission between rodent to humans is to human to humans (*ψ* = 1). Here, for *S*_*m*_(0) = 0, *R*_0_(*t*) and *S*_*m*_(*t*) crosses the threshold to epidemic region only for *σ* = 0.5 and *σ* = 0.9. But starting with *S*_*m*_(0) = 2500 and *S*_*m*_(0) = 10000, all started in the epidemic region. This is due to the high initial reproductive number and initial density of susceptibles above the critical threshold.

It is important to note that despite high *R*_0_, with low initial number of susceptibility to MPXV, no epidemic is initiated coupled with high immunity (*σ* = 0) in the population. This makes us understand that increasing the population immunity and reducing the population of susceptible can alter the risk of MPXV epidemic, especially when the potential of infection growth (*R*_0_) is high in the population.

### 4.8 Effect of Waning Immunity on Long-Term Dynamics of MPXV

The effect of waning immunity on the long-term dynamics of MPXV is illustrated in Fig. 10-12. This analysis examines various levels of *ψ* = 0, 0.5, and 1, representing low, moderate, and high transmission scenarios (*R*_0_). The initial sizes of the MPXV-susceptible population considered are *S*_*m*_(0) = 0, 2500, and 10000, while the immune compartment is initialized with a constant size of *R*_*s*_(0) = 10000. The dynamics are studied over an extended period (*t* = 200) to capture critical features, such as the impact of waning immunity on long-term epidemic behavior. This extended time frame allows for a comprehensive understanding of how waning immunity influences the progression and persistence of MPXV in different transmission scenarios.

**Fig. 10:**
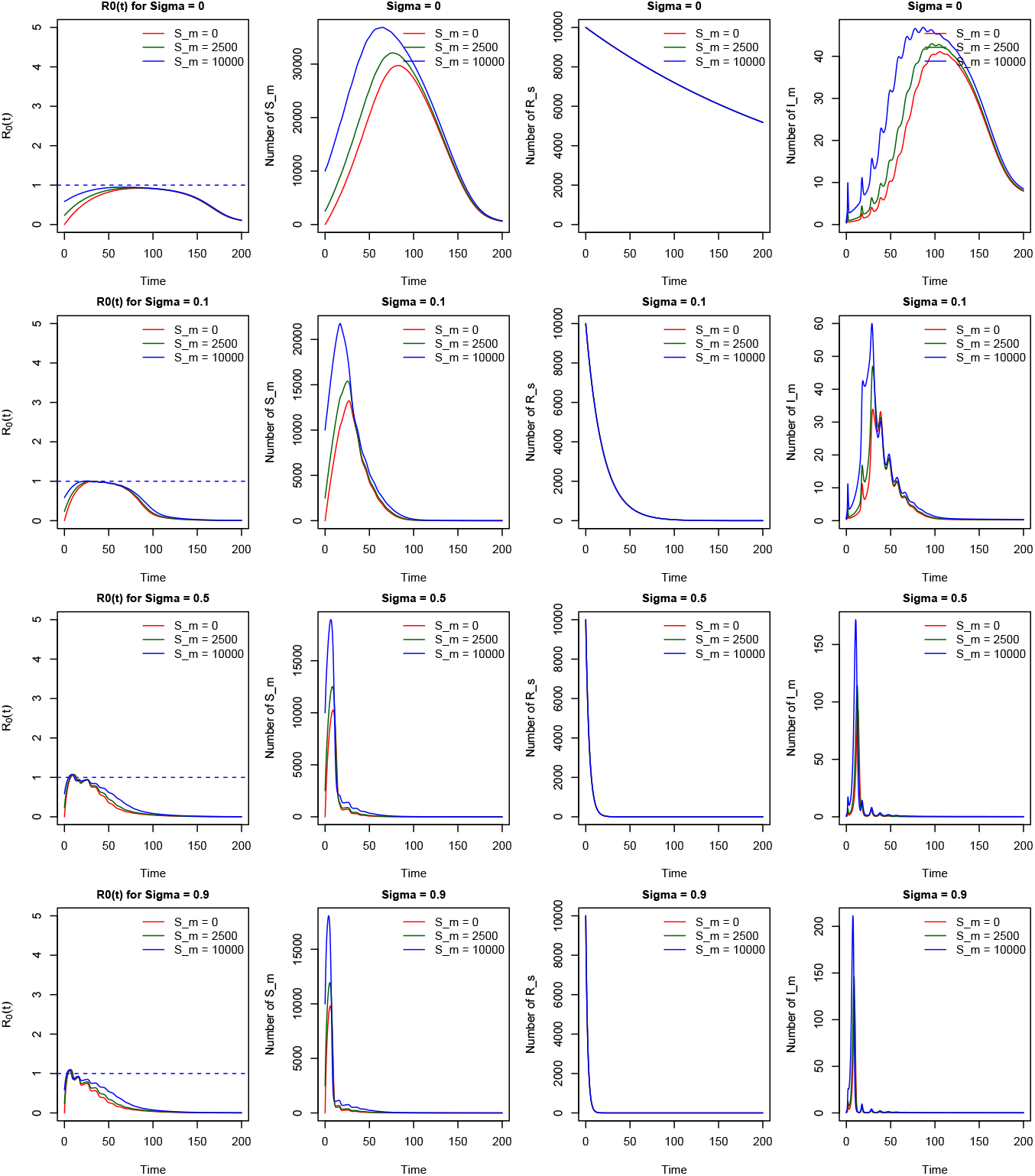
plot showing the effect of waning immunity on long term dynamics of *R*_0_(*t*), *S*_*m*_(*t*), *R*_*s*_(*t*), and *I*_*m*_(*t*) for various levels of waning immunity (*σ* = 0, 0.1, 0.5, and 0.9) for low transmission potential. The plot examines the effect of waning immunity on the threshold dynamics with initial conditions *S*_*m*_(0) = 0, 2500, and 10000 and *R*_*s*_(0) = 10000. For *R*_0_(0) < 1, the relative infectivity rate of rodent to human is *ψ* = 0. For period of T=200.

Fig. 10 above illustrates the plots of *R*_0_(*t*), *S*_*m*_(*t*), *R*_*s*_(*t*), and *I*_*m*_(*t*). No epidemic was initiated throughout the entire period when the population was assumed to have permanent immunity. This outcome is evident in the low transmission dynamics observed, as *R*_0_(*t*) ≤ 1 for all *t* ≥ 0. The curves for *R*_0_(*t*) and *I*_*m*_(*t*) further confirm the lack of epidemic activity, which can be attributed to the low transmission profile within the population.

despite increase in the waning rate from low (*σ* = 0) to high (*σ* = 0.9)waning rate, there is no change in the epidemic status. The system remains in the non-epidemic region, and the only observable changes in the dynamic behavior is on the duration times. The peak time decreases with increased waning rate.

In Fig. 11, oscillatory dynamics are observed, indicating waves of minor epidemics following the initial major epidemic. These waves are evident in the plots showing the dynamics of *R*_0_(*t*) and the profiles of *S*_*m*_(*t*) and *I*_*m*_(*t*). The amplitude of the waves decreases as the waning rate increases from low (*σ* = 0) to high (*σ* = 0.9), highlighting the impact of faster immunity loss on the epidemic patterns.

**Fig. 11:**
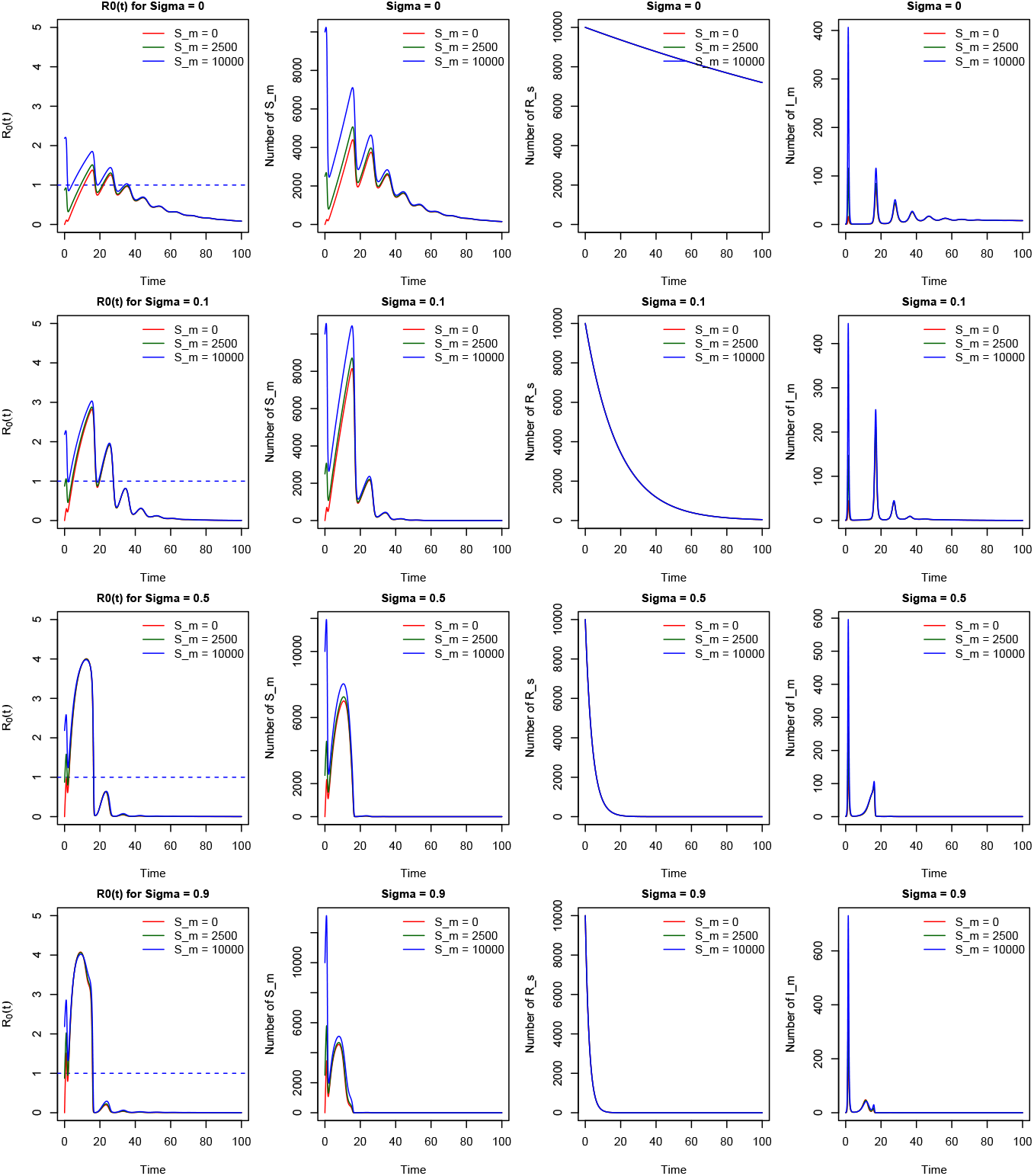
plot showing the effect of waning immunity on long term dynamics of *R*_0_(*t*), *S*_*m*_(*t*), *R*_*s*_(*t*), and *I*_*m*_(*t*) for various levels of waning immunity (*σ* = 0, 0.1, 0.5, and 0.9) for moderate transmission potential. The plot examines the effect of waning immunity on the threshold dynamics with initial conditions *S*_*m*_(0) = 0, 2500, and 10000 and *R*_*s*_(0) = 10000. For *R*_0_(0) ≥ 0.5, the relative infectivity rate of rodent to human is *ψ* = 0.5. For period of T=200.

These waves result from a slow waning rate, which causes a prolonged inter-epidemic period for each cycle. Additionally, the dynamic patterns in *R*_0_(*t*) and *S*_*m*_(*t*) remain consistent over time, highlighting the implications of the dual threshold theory. This unification is introduced through the linkage between the temporal dynamics of *R*_0_(*t*) and *S*_*m*_(*t*), as influenced by waning immunity dynamics (*σ*) and population dynamics (birth and death processes). This pattern is consistent across all initial susceptible populations (*S*_*m*_(0) = 0, 2500, and 10000). The number of cases in *I*_*m*_(*t*) increases as the waning rate (*σ*) increases. Additionally, this dynamic pattern mirrors the observations in Fig. 12, further emphasizing the relationship between waning immunity and the temporal dynamics of MPXV transmission.

**Fig. 12:**
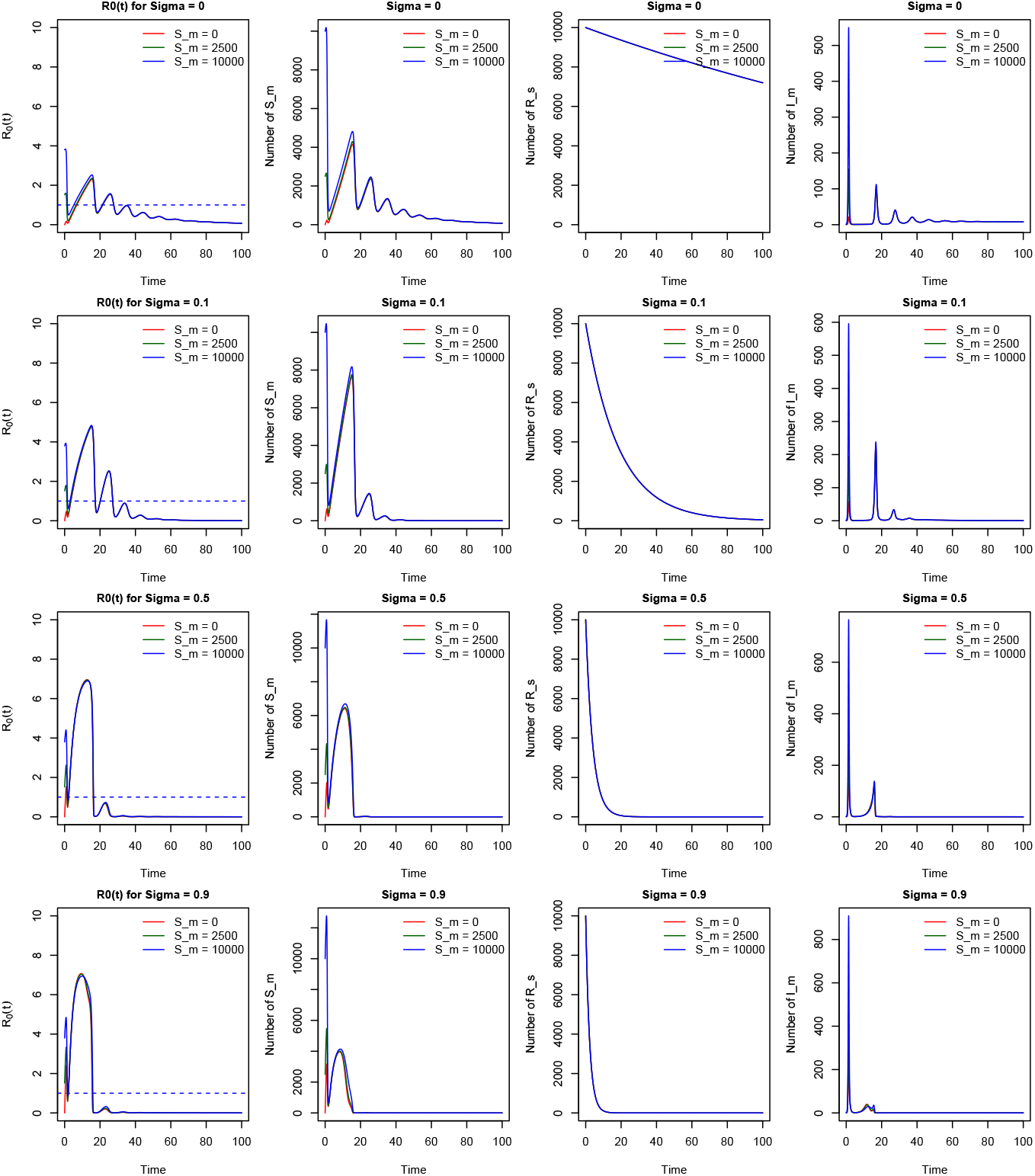
plot showing the effect of waning immunity on long term dynamics of *R*_0_(*t*), *S*_*m*_(*t*), *R*_*s*_(*t*), and *I*_*m*_(*t*) for various levels of waning immunity (*σ* = 0, 0.1, 0.5, and 0.9) for High transmission potential. The plot examines the effect of waning immunity on the threshold dynamics with initial conditions *S*_*m*_(0) = 0, 2500, and 10000 and *R*_*s*_(0) = 10000. For *R*_0_(0) ≥ 1, the relative infectivity rate of rodent to human is *ψ* = 1. For period of T=200

## 5 Constructing Epidemic Curve using Dynamic reproductive number

Constructing an epidemic curve for diseases with waning immunity, such as monkeypox and meningitis (Yaga and Saporu, 2024a), is not straightforward due to the temporal dynamics in the susceptible compartment. To account for these transitions, we derive the epidemic curve using the temporal changes in the time-dependent reproductive number, *R*_0_(*t*).

Becker (2015) constructed epidemic curves for successive generations using the SIR-type epidemic models with a static basic reproductive number. This approach can be generalized for diseases with significant temporal dynamics in the susceptible class, driven by factors such as waning immunity (from vaccination or infection) and population births.

The basic reproductive number, *R*_0_, is defined as the potential average number of new cases generated by a single infectious person in a wholly susceptible population. This definition forms the basis of our formalism.

### 5.0.1 Epidemic Curve

At time *t* = *j*, let *I*(*j*) represent the number of infectious individuals, *S*(*j*) the number of susceptibles, and *N*(*j*) the total population size. Each infectious individual produces, on average, *R*_0_ new cases during their infectious period.

When *I*(*j*) infectious individuals come into contact with a fraction of susceptibles (*S*(*j*)*/N* (*j*)), the total number of new cases produced will be:

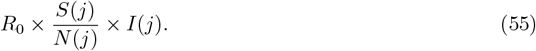

This approach can be applied iteratively to estimate the number of new cases for each successive generation, corresponding to the end of the latent period. The number of new cases generated at each time step can be expressed as:

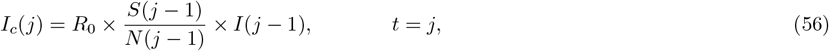

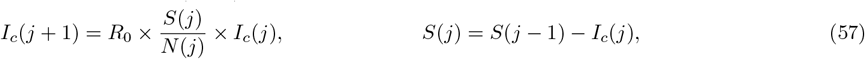

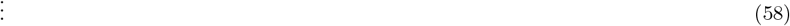

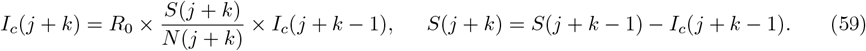

where *I*_*c*_(*j* + *k*), *k* = 0, 1, 2, …, *n*, represents the number of new cases in the *k*^*th*^ − generation, *S*(*j* + *k*) is the number of susceptibles at time *j* in generation *k, N* (*j* + *k*) is the total population size in *k*^*th*^− generation and *R*_0_ is the basic reproductive number. This iterative process provides a dynamic framework for constructing the epidemic curve, capturing the progression of new cases over time. It accounts for the depletion of susceptibles and changes in population size, enabling the generation of an epidemic curve that reflects real-world dynamics.

### 5.1 Role of Waning Immunity in Shaping Epidemic Curves

The dynamics of *R*_0_(*t*) enable us to understand the intrinsic salient features of waning immunity on the dynamics of epidemic transmissions across the epidemic and non-epidemic regions.

Epidemic curves illustrate the incidence dynamics under varying levels of waning immunity (*σ*, *σ*_*m*_), studied across three scenarios representing low, moderate, and high transmission potentials at the initial time (*R*_0_ at *t* = 0), as shown in Fig. 13-15. For low transmission (Fig. 13), slower replenishment of susceptibles leads to a rapid depletion of susceptibles during the epidemic peak, which limits sustained transmission. This causes a steeper decline and earlier termination of the epidemic, as the population quickly reaches the herd immunity threshold. The low transmission scenario is characterized by low intensity, producing smaller, flatter peaks due to reduced transmission efficiency, with a longer time to peak compared to scenarios with higher transmission, as observed in Fig. 14 and Fig. 15, where the epidemic grows more gradually.

**Fig. 13:**
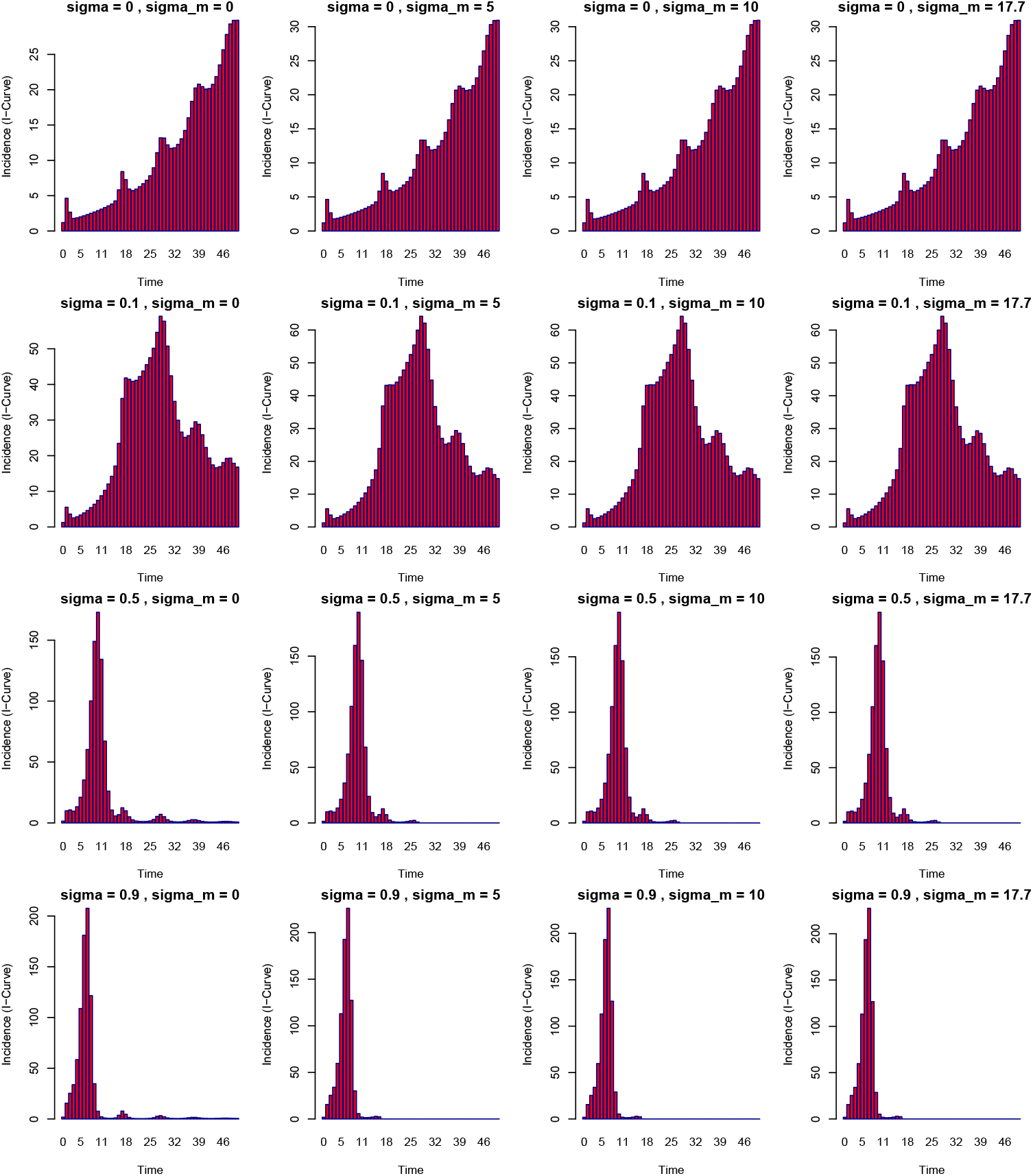
Showing the incidence dynamics under varying levels of waning immunity (*σ* = 0, 0.1, 0.5 and *σ* = 0.9) for different values of *σ*_*m*_. The plot examines the effect of waning immunity on the shapes of epidemic curves for low transmission potential with initial conditions *S*_*m*_(0) = 10000 and *R*_*s*_(0) = 10000,*I*_*m*_(0) = 1), the relative infectivity rate of rodent to human is *ψ* = 0

**Fig. 14:**
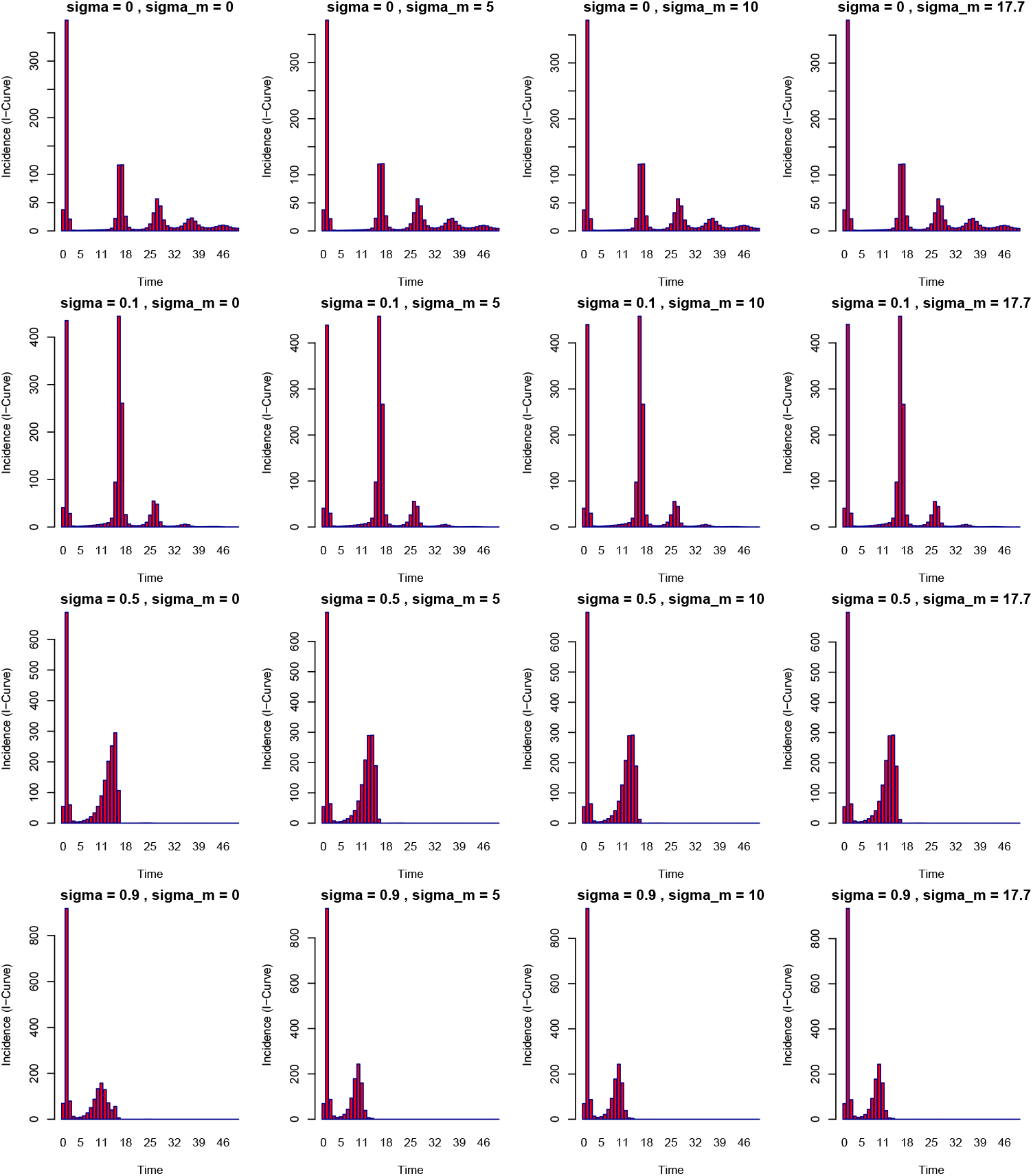
Showing the incidence dynamics under varying levels of waning immunity (*σ* = 0, 0.1, 0.5 and *σ* = 0.9) for different values of *σ*_*m*_. The plot examines the effect of waning immunity on the shapes of epidemic curves for moderate transmission potential with initial conditions *S*_*m*_(0) = 10000 and *R*_*s*_(0) = 10000,*I*_*m*_(0) = 1), the relative infectivity rate of rodent to human is *ψ* = 0.5

**Fig. 15:**
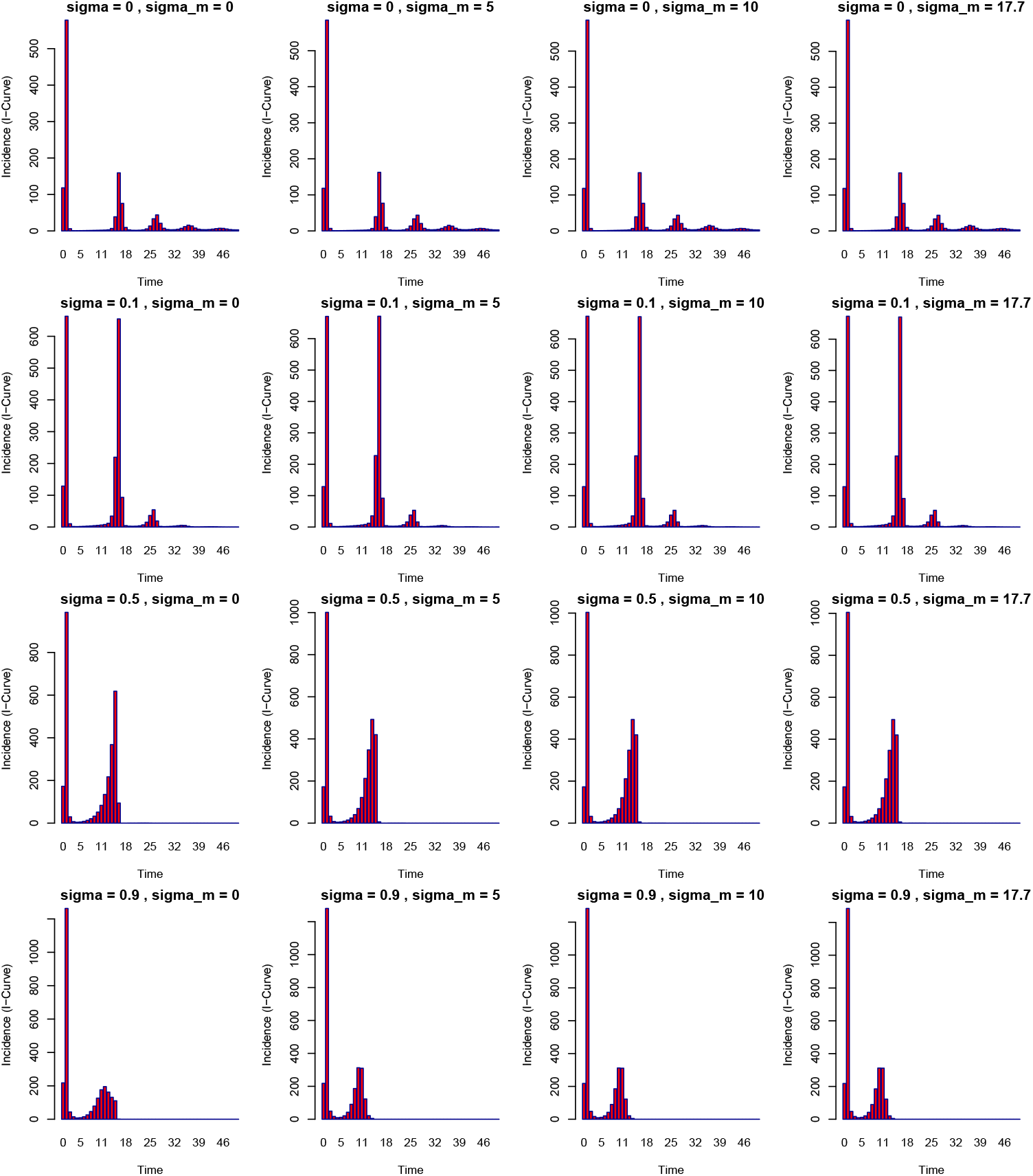
Showing the incidence dynamics under varying levels of waning immunity (*σ* = 0, 0.1, 0.5 and *σ* = 0.9) for different values of *σ*_*m*_. The plot examines the effect of waning immunity on the shapes of epidemic curves for High transmission potential with initial conditions *S*_*m*_(0) = 10000 and *R*_*s*_(0) = 10000,*I*_*m*_(0) = 1), the relative infectivity rate of rodent to human is *ψ* = 1

For low waning rates (*σ* = 0, 0.1), epidemics exhibit smaller, more symmetric peaks, indicating a balanced rise and fall in cases. Higher waning rates, however, lead to slightly taller and flatter peaks, especially in populations with high transmission potential (*R*_0_(*t*)). Fig. 13 highlights that slower waning rates prolong epidemics by gradually depleting susceptibles, while rapid waning rates sustain transmission for longer due to faster replenishment of susceptibles. Epidemic intensity is lower under low transmission potential (*R*_0_(*t*)) compared to high transmission potential, as shown in Fig. 13-15. In moderate and high transmission scenarios (Fig. 14 and 15), epidemics feature steep, narrow peaks with rapid case surges, especially when waning rates are low. This underscores the interplay between waning immunity and transmission potential in shaping epidemic dynamics and case burden.

#### 5.1.1 Impact of Waning Immunity and Rodent Infectivity on Epidemic profiles

Table 4 is computed from Figures 16-18. we are interested in exploring the relationship between peak times (*T*_*p*_) and peak cases (*I*_*c*_(*T*_*p*_)) across various levels of waning immunity rates (*σ*) and mixing parameters (*σ*_*m*_), alongside varying values of the rodent infectivity rate (*ψ*)—which takes values of 0, 0.5, and 1. This setup provides insights into the dynamics of infectious disease spread, particularly how these parameters interact to influence epidemic trajectories.

**Table 4:** Peak Times (*T*_*p*_) and Peak Cases (*I*_*c*_(*T*_*p*_)) Across Waning Immunity Rates (*σ*) and Mixing Parameters (*σ*_*m*_) for Various *ψ* Values (0, 0.5, 1)

| Waning Immunity | | $\psi = 0$ | | $\psi = 0.5$ | | $\psi = 1$ | |
| --- | --- | --- | --- | --- | --- | --- | --- |
| $\sigma$ | $\sigma_m$ | Peak Time ($T_p$) | Peak Cases $I_c(T_p)$ | Peak Time ($T_p$) | Peak Cases $I_c(T_p)$ | Peak Time ($T_p$) | Peak Cases $I_c(T_p)$ |
| 0.0 | 0 | 50.000 | 27.048 | 1.4710 | 295.407 | 1.370 | 611.153 |
| 0.0 | 5 | 50.000 | 28.029 | 1.4740 | 296.445 | 1.370 | 613.765 |
| 0.0 | 10 | 50.000 | 28.034 | 1.4740 | 296.768 | 1.370 | 614.628 |
| 0.0 | 17 | 50.000 | 28.036 | 1.4740 | 296.972 | 1.370 | 615.191 |
| 0.1 | 0 | 26.312 | 58.489 | 17.134 | 429.391 | 1.375 | 707.894 |
| 0.1 | 5 | 26.436 | 64.893 | 17.137 | 442.635 | 1.378 | 711.072 |
| 0.1 | 10 | 26.433 | 64.978 | 17.137 | 443.154 | 1.378 | 712.135 |
| 0.1 | 17 | 26.430 | 64.893 | 17.14 | 443.419 | 1.378 | 712.827 |
| 0.5 | 0 | 10.353 | 178.415 | 1.5150 | 580.108 | 1.389 | 1115.278 |
| 0.5 | 5 | 10.408 | 194.775 | 1.5180 | 582.909 | 1.392 | 1121.097 |
| 0.5 | 10 | 10.408 | 195.568 | 1.5180 | 583.790 | 1.392 | 1123.075 |
| 0.5 | 17 | 10.408 | 195.930 | 1.5180 | 584.341 | 1.392 | 1124.368 |
| 0.9 | 0 | 7.5590 | 243.347 | 1.5290 | 823.868 | 1.389 | 1518.291 |
| 0.9 | 5 | 7.6000 | 261.814 | 1.5320 | 827.868 | 1.392 | 1527.003 |
| 0.9 | 10 | 7.6030 | 263.036 | 1.5320 | 829.368 | 1.392 | 1529.987 |
| 0.9 | 17 | 7.6030 | 262.807 | 1.5320 | 830.305 | 1.392 | 1531.947 |

**Fig. 16:**
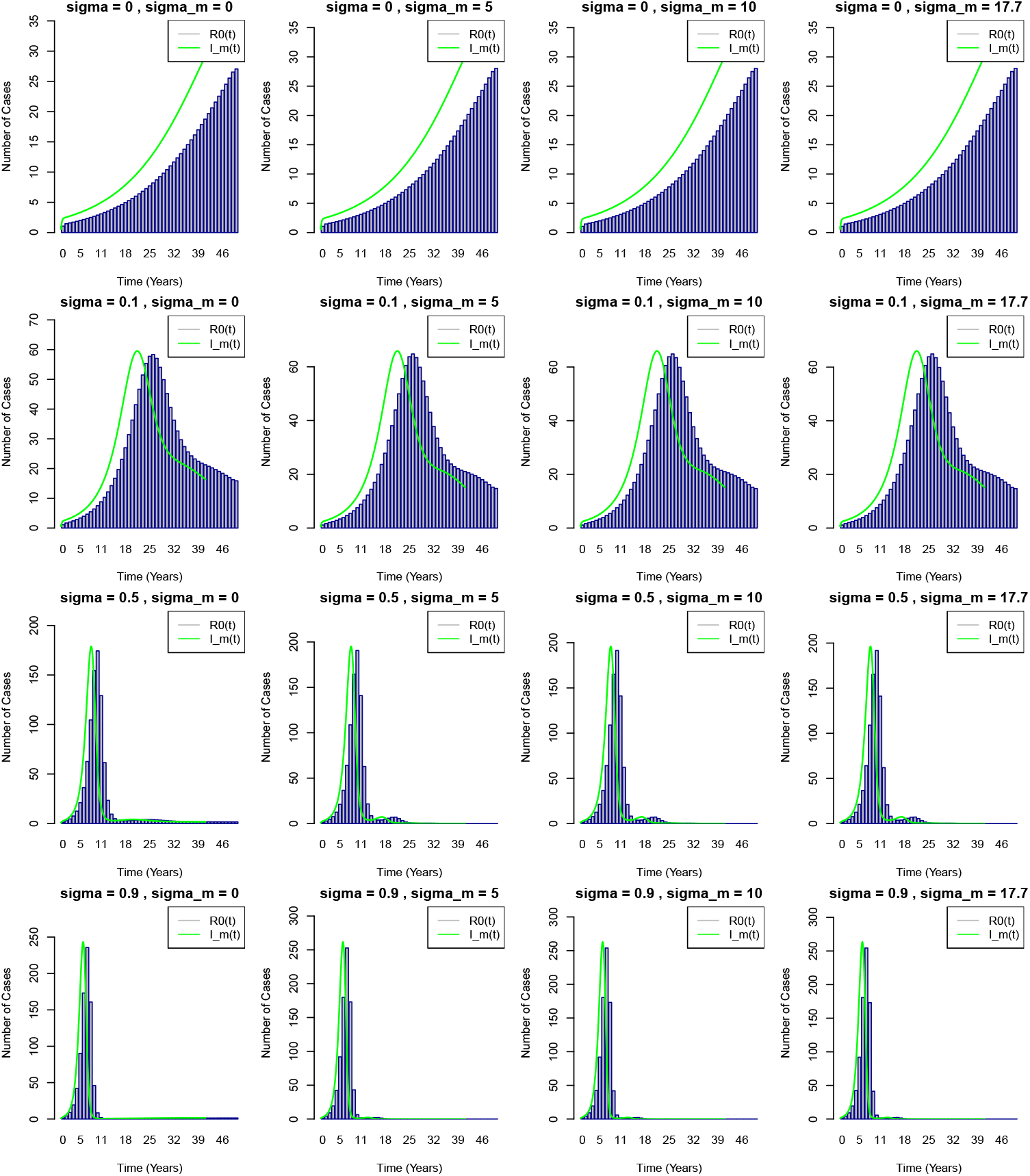
Incidence Curves for varying levels of waning immunity (*σ* = 0, 0.1, 0.5 and *σ* = 0.9) for different values of *σ*_*m*_. The plot examines the epidemiological characteristics of epidemic incidence such as peak time and number of cases at peak time, presented in Table(4) for low transmission potential with initial conditions *S*_*m*_(0) = 10000 and *R*_*s*_(0) = 10000,*I*_*m*_(0) = 1), the relative infectivity rate of rodent to human is *ψ* = 0

**Fig. 17:**
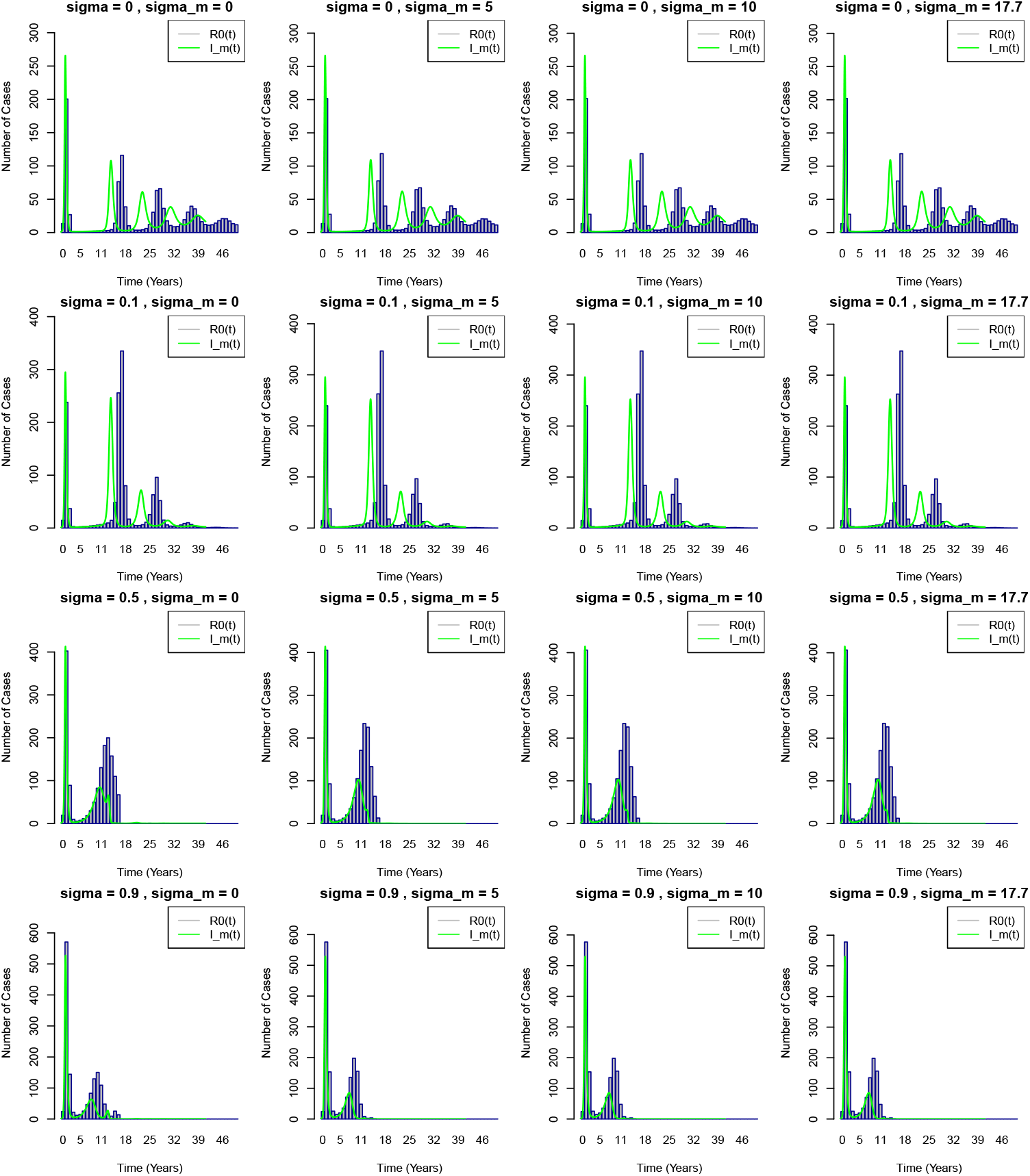
Incidence Curves for varying levels of waning immunity (*σ* = 0, 0.1, 0.5 and *σ* = 0.9) for different values of *σ*_*m*_. The plot examines the epidemiological characteristics of epidemic incidence such as peak time and number of cases at peak time, presented in Table(4) for moderate transmission potential with initial conditions *S*_*m*_(0) = 10000 and *R*_*s*_(0) = 10000,*I*_*m*_(0) = 1), the relative infectivity rate of rodent to human is *ψ* = 0.5

**Fig. 18:**
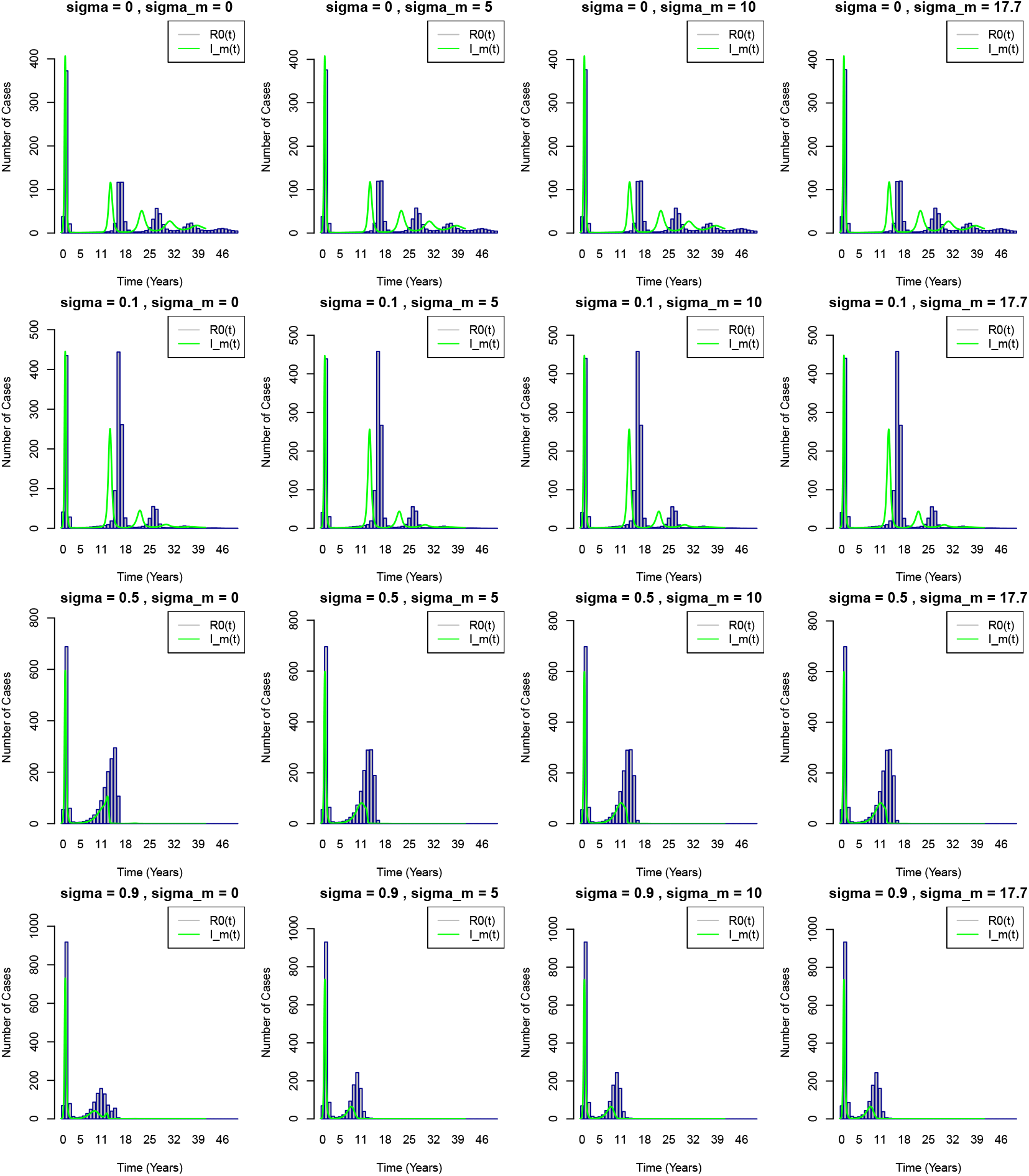
Incidence Curves for varying levels of waning immunity (*σ* = 0, 0.1, 0.5 and *σ* = 0.9) for different values of *σ*_*m*_. The plot examines the epidemiological characteristics of epidemic incidence such as peak time and number of cases at peak time, presented in Table(4) for High transmission potential with initial conditions *S*_*m*_(0) = 10000 and *R*_*s*_(0) = 10000,*I*_*m*_(0) = 1), the relative infectivity rate of rodent to human is *ψ* = 1

In the context of Table 4, we observe that peak times (*T*_*p*_) and peak cases (*I*_*c*_(*T*_*p*_)) exhibit distinct patterns based on varying values of waning immunity rates (*σ*), rodent infectivity rates (*ψ*), and immunity loss from previous monkeypox infection (*s*_*m*_).

1. **Waning Immunity (***σ*): The rate at which immunity declines from vaccination (*σ*) plays a crucial role in determining both the timing and severity of the epidemic. As *σ* increases, we observe a shift in the peak time to earlier time points and an increase in the peak cases. For example, when *σ* = 0.0, the peak time is at 50 days across all mixing parameters (*σ*_*m*_) and the peak cases increase as *ψ* increases from 0 to 1. However, as *σ* rises, the peak time decreases (e.g., at *σ* = 0.9, peak times are reduced to around 7.6 days), indicating faster epidemic dynamics, along with higher peak cases, especially in the higher values of *ψ*.
2. **Rodent Infectivity (***ψ*): The impact of the rodent infectivity rate (*ψ*) is significant across all values of *σ* and *σ*_*m*_. For example, at lower values of *ψ* (e.g., *ψ* = 0), the peak times are longer, and the peak cases are lower. As *ψ* increases, the dynamics shift towards earlier peak times and larger peak cases. This reflects the increased transmission potential from rodents, which accelerates the overall epidemic dynamics and results in more cases.
3. **Immunity Loss from Recovery (***σ*_*m*_): The parameter *σ*_*m*_ affects the immunity loss from individuals who have recovered from monkeypox infection. This factor does not have a major impact on the peak time; however, it does influence the number of peak cases. As *σ*_*m*_ increases, there is a slight increase in peak cases, particularly noticeable at higher values of *ψ*, indicating that faster loss of immunity from recovered individuals may contribute to the higher number of susceptible individuals, enhancing epidemic spread.

The results presented in the table have several important epidemiological implications, particularly for the control and prediction of future outbreaks. For vaccination programs, these findings suggest that maintaining high levels of immunity in the population is crucial for controlling outbreaks. If immunity from vaccination wanes too quickly, individuals may become susceptible again, which can lead to resurgence of cases. Regular booster doses or new vaccination strategies may be required to maintain sufficient herd immunity levels and delay or prevent peak outbreaks.

In conclusion, the table suggests a complex interplay between waning immunity, rodent infectivity, and immunity loss from recovery, each influencing the peak dynamics of the epidemic. The faster waning immunity and higher rodent infectivity increase the epidemic’s severity and speed. Understanding these relationships is crucial for developing strategies to mitigate future outbreaks, especially by addressing vaccination policies, rodent control measures, and post-infection immunity recovery.

## 6 Conclusion

This study advances epidemic theory by introducing the dual threshold framework, offering a novel perspective on Monkeypox virus (MPXV) transmission dynamics. It formalizes the concept of epidemic regions, determined by the critical interaction between the time-dependent reproductive number (*R*_0_(*t*)) and the susceptible population density (*S*_*m*_(*t*)). The dual threshold theory establishes that epidemic initiation and progression require the system to cross two boundaries: *R*_0_(*t*) > 1 and *S*_*m*_(*t*) > *S*_*c*_(*t*), where *S*_*c*_(*t*) represents the critical density of susceptibles necessary to sustain transmission.

This framework provides a deeper understanding of the transition between epidemic and non-epidemic regions, highlighting the interdependence between *R*_0_(*t*) and *S*_*m*_(*t*). Notably, *R*_0_(*t*) > 1 alone is insufficient to sustain transmission unless *S*_*m*_(*t*) also exceeds *S*_*c*_(*t*), underscoring the importance of jointly monitoring these thresholds.

The study further examines the role of waning immunity, including immunity loss due to smallpox vaccination (*σ*) and prior MPXV infections (*σ*_*m*_). The findings reveal complex epidemic dynamics, such as oscillatory waves, prolonged inter-epidemic periods, and varying epidemic intensities across different transmission scenarios. These results highlight the significant influence of waning immunity in shaping both short-term and long-term epidemic trajectories.

1. **Impact of Waning Immunity**: Faster waning rates (*σ*, *σ*_*m*_) accelerate epidemic cycles, increasing peak cases and shortening inter-epidemic periods. These dynamics emphasize the necessity of robust immunization strategies, including regular booster campaigns, to sustain population immunity.
2. **Role of Zoonotic Reservoirs**: Rodent infectivity (*ψ*) plays a critical role in sustaining transmission, particularly under high transmission potential. Public health measures focusing on rodent control are essential to mitigate spillover risks and reduce epidemic severity.
3. **Temporal Dynamics**: Oscillatory patterns in *R*_0_(*t*), *S*_*m*_(*t*), and *I*_*m*_(*t*) under moderate and high transmission scenarios underscore the intricate relationship between population susceptibility and epidemic peaks. This calls for adaptive public health strategies to address shifting epidemic dynamics.
4. **Public Health Implications**: Continuous monitoring of *R*_0_(*t*) and *S*_*m*_(*t*) provides early warning signals for potential outbreaks. Integrative approaches combining vaccination campaigns, rodent control, and surveillance are vital to managing MPXV dynamics and preventing resurgence.

This study unifies threshold-based epidemic theory with dynamic population modeling, offering a comprehensive framework for understanding MPXV transmission. By delineating epidemic and non-epidemic regions, the dual threshold theory provides a foundation for future research on zoonotic diseases and informs practical strategies for epidemic management.

By formalizing the conditions under which epidemics are sustained or mitigated, this research contributes significantly to theoretical epidemiology and provides actionable guidance for managing MPXV outbreaks. The findings emphasize the necessity of integrated One Health approaches that account for immunity dynamics, population thresholds, and zoonotic reservoirs within environmental contexts to prevent and control future epidemics. This study not only enhances theoretical models of zoonotic disease dynamics but also underscores the critical importance of maintaining immunity in at risk populations and addressing zoonotic reservoirs to reduce the risk of MPXV and other emerging diseases.

## Data Availability

All data produced in the present work are contained in the manuscript

## Notes

### Competing Interest Statement

The authors have declared no competing interest.

### Funding Statement

This study did not receive any funding

### Author Declarations

My research does not involve the use of human data or human subjects. It focuses on theoretical modeling and mathematical analysis of disease dynamics, particularly the impact of waning immunity from smallpox vaccination on monkeypox transmission. There is no collection, analysis, or use of human data, patient records, or biological samples

